# Action-sequence learning, habits and automaticity in obsessive-compulsive disorder

**DOI:** 10.1101/2023.02.23.23286338

**Authors:** Paula Banca, Maria Herrojo Ruiz, Miguel Fernando Gonzalez-Zalba, Marjan Biria, Aleya A. Marzuki, Thomas Piercy, Akeem Sule, Naomi Anne Fineberg, Trevor William Robbins

## Abstract

Enhanced habit formation, greater automaticity and impaired goal/habit arbitration in obsessive-compulsive disorder (OCD) are key hypotheses from the goal/habit imbalance theory of compulsion which have not been directly investigated. This study tests these hypotheses using a combination of newly developed behavioral tasks. First, we trained both OCD patients and healthy controls, using a smartphone app, to perform chunked action sequences. This motor training was conducted daily for one month. Both groups displayed equivalent procedural learning and attainment of habitual performance (measured with an objective criterion of automaticity), despite greater subjective habitual tendencies in patients with OCD, self-reported via a recently developed questionnaire. Participants were subsequently tested on a re-evaluation task to assess choice between established automatic and novel goal-directed action sequences. This task showed that both groups were sensitive to re-evaluation based on monetary feedback. However, when re-evaluation was based on physical effort, OCD patients showed a pronounced preference for the previously trained habitual sequence, hypothetically due to its intrinsic value. This was particularly evident in patients with higher compulsive symptoms and habitual tendencies, who also engaged significantly more with the motor habit-training app and reported symptom relief at the end of the study. The tendency to attribute higher intrinsic value to familiar actions may be a potential mechanism leading to compulsions and an important addition to the goal/habit imbalance hypothesis in OCD. We also highlight the potential of the app-training as a habit reversal therapeutic tool.

## Introduction

Considerable evidence has supported the concept of imbalanced cortico-striatal pathways mediating compulsive behavior in obsessive-compulsive disorder (OCD). This imbalance has been suggested to reflect a weaker goal-directed control and an excessive habitual control (Gillan et al., 2016). Dysfunctional goal-directed control in OCD has been strongly supported both behaviorally (Gillan et al., 2011; Vaghi et al., 2018) and from a neurobiological perspective (Gillan et al., 2015a). However, until now, enhanced (and potentially maladaptive) habit formation has largely been inferred by the absence of goal-directed control, although recent studies show increased self-reported habitual tendencies in OCD, as measured by the Self-Report Habit Index Scale (Ferreira et al., 2017). Problems with this “zero-sum” hypothesis (Robbins and Costa, 2017) (i.e., diminished goal directed control *thus* enhanced habitual control) have been underlined by recent findings linking stimulus-response strength (Zwosta et al., 2018) and goal devaluation (Gillan et al., 2015b) exclusively to a dysfunctional goal system. There is thus a need to focus specifically on the habit component of the associative dual-process (i.e. goal/habit) model of behavior and test more directly the hypothesis of enhanced habit formation in OCD.

We recently proposed that extensive training of sequential actions could be a means for rapidly engaging the ‘habit system’ in a laboratory setting (Robbins et al., 2019). The idea is that, in action sequences (like those seen in skilled routines), extensive training helps integrate separate motor actions into a coordinated and unified sequence, or “chunk” (Graybiel, 1998; Sakai et al., 2003). Through consistent practice, the selection and execution of these component actions become more streamlined, stereotypical, and cognitively effortless. They are performed with minimal variation, achieving high efficiency. Moreover, there is now robust evidence that for highly-trained sequences, actions are represented in parallel according to their serial order before execution (Kornysheva et al., 2019). Such features relate to the concept of *automaticity*, which captures many of the shared elements between habits and skills (Ashby et al., 2010). At a neural level, automaticity is associated with a shift in control from the anterior/associative (goal-directed) to the posterior/sensorimotor (habitual) striatal regions (Ashby et al., 2010; Graybiel and Grafton, 2015; Kupferschmidt et al., 2017), accompanied by a disengagement of cognitive control hubs in frontal and cingulate cortices (Bassett et al., 2015). In fact, within the skill learning literature, this progressive shift to posterior striatum has been linked to the gradually attained asymptotic performance of the skill (Bassett et al., 2015; Doyon et al., 2018, 2015; Lehericy et al., 2005). Hence chunked action sequences provide an opportunity to target the brain’s goal-habit transition and study the relationships between automaticity, skills and habits (Dezfouli et al., 2014; Graybiel and Grafton, 2015; Robbins and Costa, 2017). This approach is relevant for OCD research as it mimics the sequences of motor events and routines observed in typical compulsions, often performed in a ‘‘just right’’ manner (Hellriegel et al., 2017), akin to skill learning. Chunked action sequences also enable investigation of the relationships between hypothesized procedural learning deficits in OCD (Rauch et al., 1997) and automaticity.

Following this reasoning, we developed a smartphone *Motor Sequencing App* with attractive sensory features in a game-like setting, to investigate automaticity and measure habit/skill formation within a naturalistic setting (at home). This task, akin to a piano-based app, allows subjects to learn and practice two sequences of finger movements. It was tailored to emphasize the positive aspects of habits, as advocated by Watson et al., 2022, and satisfies central criteria that define habits proposed by Balleine and Dezfouli, 2019: swift execution, invariant response topography and action chunking. We also aimed to investigate within the same experiment three facets of automaticity which, according to Haith and Krakauer (2018), have rarely been measured together: habit, skill and cognitive load. Although there is no consensus on how exactly skills and habits interact (Robbins and Costa, 2017), it is generally agreed that both lead to automaticity with sufficient practice (Graybiel and Grafton, 2015) and that the autonomous nature of habits and the fluid proficiency of skills engage the same sensorimotor cortical-striatal ‘loops’ (the so-called ‘habit circuitry’) (Ashby et al., 2010; Graybiel and Grafton, 2015). By focusing more on the *automaticity* of the response per se (as reflecting the speed and stereotypy of over-trained movement sequences), rather than on the *autonomous* nature of the behavior (an action that continues after a state change, e.g. devaluation of the goal), we do not solely rely on the devaluation criterion used in previous studies of compulsive behavior. This is important because outcome devaluation insensitivity as a test of habit in humans is controversial (Watson et al., 2022) and may indeed be a more sensitive indicator of failures of goal-directed control rather than of habitual control per se (Balleine and Dezfouli, 2019; Robbins et al., 2019; Robbins and Costa, 2017).

While designing our app, we additionally considered previous research emphasizing training frequency, context stability, and reward contingencies as important features for enhancing habit strength (Wood and Rünger, 2016). To ensure effective consolidation required for habit/skill retention to occur, we implemented a 1-month training period. This aligns with studies showing that practice alone is insufficient for habit development as it also requires off-line consolidation over longer periods of time and sleep (Nusbaum et al., 2018; Walker et al., 2003). Finally, given the influence of reinforcer predictability on habit acquisition speed (Bouton, 2021) we employed two different reinforcement schedules (reward scores: continuous versus variable [probabilistic]) to assess their impact on habit formation amongst healthy volunteers (HV) and patients with OCD.

### Outline

In this article, we applied, for the first time, an app-based behavioral training (experiment 1) to a sample of patients with OCD. We compared 32 patients and 33 healthy participants, matched for age, gender, IQ and years of education in measures of motivation and app engagement (see Methods for participants’ demographics and clinical characteristics). We also assessed to what extent performing such repetitive actions in one month impacted OCD symptomatology. In an *initial phase* (30 days), two action sequences were trained daily to produce habits/automatic actions (experiment 1). We collected data online continuously to monitor engagement and performance in real-time. This approach ensured we acquired sufficient data for subsequent analysis of procedural learning and automaticity development.

In a *second phase,* we administered two follow-up behavioral tasks (experiments 2 and 3) addressing two important questions relevant to the habit theory of OCD. The first research question investigated whether repeated performance of motor sequences could develop implicit rewarding properties, hence gaining value, potentially leading to compulsive-like behaviors (experiment 2: explicit preference task, conducted without feedback). The hypothesis postulates that the repeated performance, initially driven by the goal of proficiency, may eventually become motivated by its own implicit reward, tied to proprioceptive and kinesthetic feedback (e.g., offering anxiety relief alongside skillful execution). The second question explored whether manipulations of extrinsic feedback, based on monetary reward or on the physical effort required (by varying the length of the sequence) affected choice for the familiar trained action sequence (experiment 3: re-evaluation task, conducted with feedback).

Finally, we administered a comprehensive set of self-reported clinical questionnaires, including a recently-developed questionnaire (Ersche et al., 2017) on habit-related aspects. This aimed to investigate: 1) if OCD patients report more habits; 2) whether stronger subjective habitual tendencies predict enhanced procedural learning, automaticity development, and an (in)ability to adjust to changing circumstances; and 3)) if app-based habit reversal therapy yields therapeutic benefits or has any subjective sequelae in OCD.

### Hypothesis

Anticipating implicit learning issues in OCD (Deckersbach et al., 2002; Kathmann et al., 2005; Rauch et al., 1997) and fine-motor difficulties (Bloch et al., 2011), we expected poorer procedural learning in patients compared to healthy volunteers. However, once learned, we predicted OCD patients would reach automaticity faster, possibly due to a stronger tendency to form habitual/automatic actions (Gillan et al., 2016, 2014). We also hypothesized differences in the learning rate and automaticity development between the two action sequences based on their associated 1) *reward schedule* (continuous versus variable), with faster automaticity in the continuous reward sequence, as suggested by past research (Bouton, 2021); and 2) *reward valence* (positive or negative), expecting enhanced performance improvements following negative feedback for all participants, especially pronounced in OCD patients due to heightened sensitivity to negative feedback (Apergis-Schoute et al 2023, Becker et al., 2014; Kanen et al., 2019). Additionally, we predicted that OCD patients would generally display stronger habits and assign greater intrinsic value to the familiar app sequences, evidenced by a marked preference for executing them even when presented with a simpler alternative sequences. Finally, we expected patients to show a greater tendency to perform the familiar/trained sequences, even though its extrinsic relative value was reduced and new, more valuable, options became available.

## Results

### Self-reported habit tendencies

Participants completed self-reported questionnaires measuring various psychological constructs (see Methods). Highly relevant for the current topic is the Creature of Habit (COHS) Scale (Ersche et al., 2017), recently developed to measure individual differences in *routine* behavior and *automatic* responses in everyday life. As compared to healthy controls, OCD patients reported significantly higher habitual tendencies both in the routine (*t* = -2.79, *p* = 0.01; 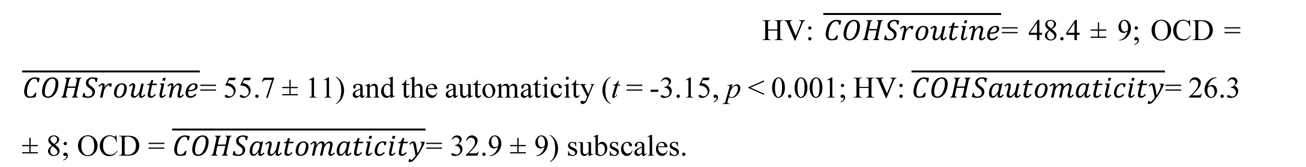

### Phase A: Experiment 1

#### Motor Sequence Acquisition using the App

The task was a self-instructed and self-paced smartphone application (app) downloaded to participants’ iPhones. It consisted of a motor practice program that participants committed to pursue daily, for a period of one month. An exhaustive description of the method has been previously published (Banca et al., 2020) but a succinct description can be found below, in Figure 1 and in the following video https://www.youtube.com/watch?v=XSYrBzD7ZpI.

**Figure 1.**
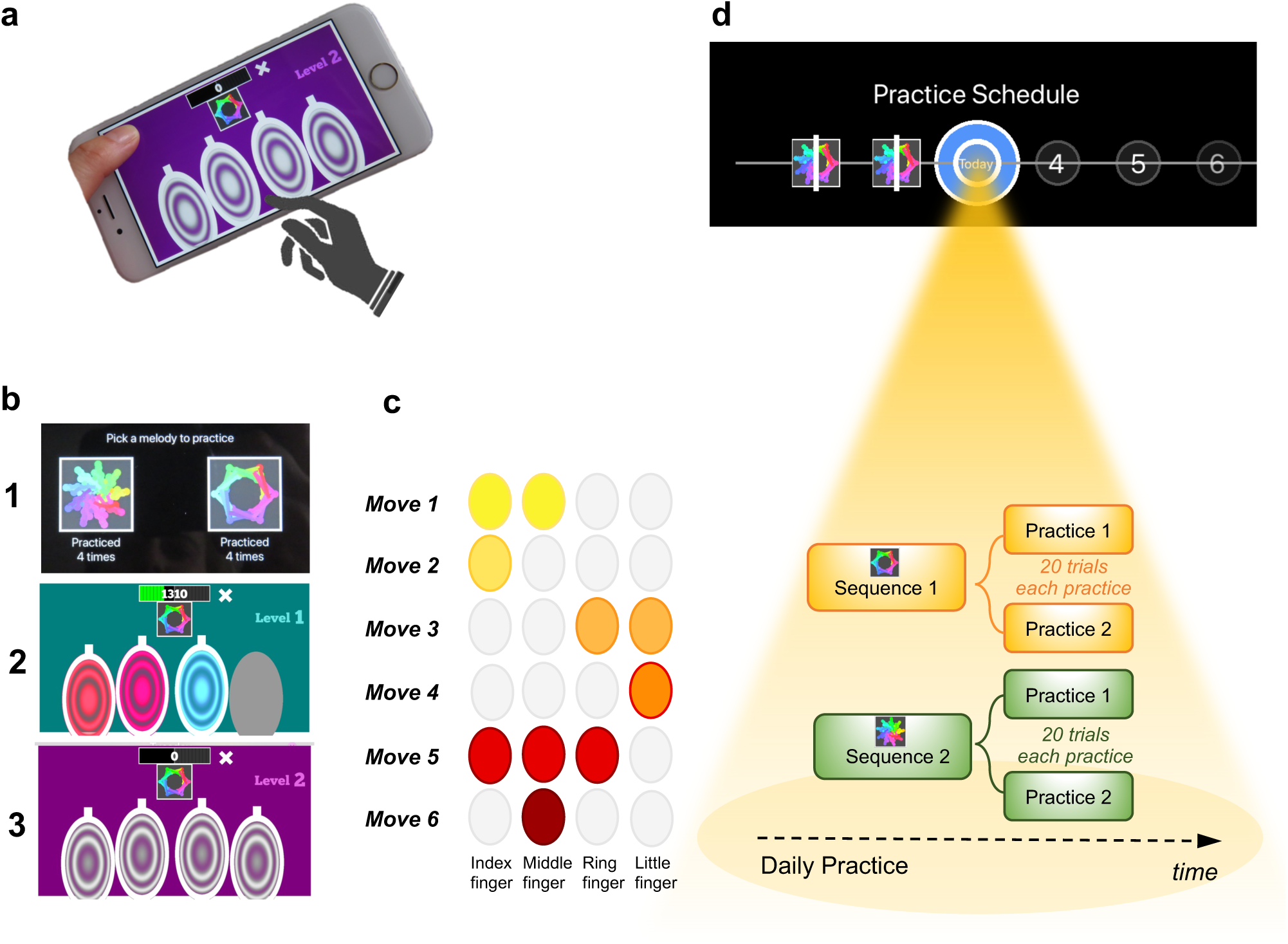
Motor Sequencing App. **a)** A trial starts with a static image depicting the abstract picture that identifies the sequence to be performed (or ’played’) as well as the 4 keys that will need to be tapped. Participants use their dominant hand to play the required keys: excluding the thumb, the leftmost finger corresponds to the first circle and the rightmost finger corresponds to the last circle. **b)** Screenshot examples of the task design: (1) sequence selection panel, each sequence is identified by an abstract picture; (2) panel exemplifying visual cues that initially guide the sequence learning; (3) panel exemplifying the removal of the visual cues, when sequence learning is only guided by auditory cues. **c)** Example of a sequence performed with the right hand: 6-moves in length, each move can comprise multiple finger presses (2 or 3 simultaneous) or a single finger press. Each sequence comprises 3 single press moves, 2 two-finger moves, and 1 three-finger move. **d)** Short description of the daily practice schedule. Each day, participants are required to play *a minimum* of 2 practices per sequence. Each practice comprised 20 successful trials. Participants could play more if they wished and the order of the training practices was self-determined.

The training consisted of practicing two sequences of finger movements, composed of chords (two or three simultaneous finger presses) and single presses (one finger only). Each sequence comprised six moves and was performed using four fingers of the dominant hand (index, middle, ring and little finger). Participants received feedback on each sequence performance (trial). Successful trials [to which we later refer as sequence trial number (*n*)] were followed by a positive ring tone and a positive visual effect (rewarding stars) and the unsuccessful ones by a negative ring tone and a negative visual effect (red lines on the screen). Every time a mistake occurred (irrespective of which move in the sequence it occurred), participants were prompted to restart the trial. Instructions were to respond swiftly and accurately. Participants were required to keep their fingers very close to the keys to minimize movement amplitude variation and to facilitate fast performance. To promote sequence learning and memorization, we implemented three progressively challenging practice levels. Initially (first 3 practice sessions), subjects responded to visual and auditory cues, following lighted keys associated with musical notes (level 1). As practice advanced, to enable motor independence and automaticity, these external cues were gradually removed: level 2 included only auditory cues (practices 4 and 5), and level 3 had no cues (remaining practices). Successful performance at each difficulty level resulted in progression to the next one. Unsuccessful performance led to reverting to the prior stage.

Each sequence, identified by a specific abstract image, was associated with a particular reward schedule. Points were calculated as a function of the time taken to complete a sequence trial. Accordingly, performance time was the instructed task-related dimension (i.e. associated with reward). In the *continuous reward schedule,* points were received for every successful trial whereas in the *variable reward schedule,* points were shown only on 37% of the trials. The rationale for having two distinct reward schedules was to assess their possible dissociable effect on the participants’ development of automatic actions. For each rewarded trial, participants could see their achieved points on the trial. To increase motivation, the total points achieved on each training session (i.e. practice) were also shown, so participants could see how well they improved across practice and days. The permanent accessibility of the app (given that most people carry their mobile phones everywhere) facilitated training frequency and enabled context stability.

##### Practice Schedule

All participants were presented with a calendar schedule and were asked to practice both sequences daily. They were instructed to practice as many times as they wished, whenever they wanted during the day and with the sequence order they would prefer. However, a minimum of 2 practices (*P*) per sequence was required every day; each practice comprised 20 successful sequence trials. Participants had to make up for missed training by completing both the current day’s session and the previous day’s if they skipped a day. If they missed training for over 2 days, the researcher gauged their motivation and incentivized their commitment. Participants were excluded if they missed training for more than 5 consecutive days.

A minimum of 30 days of training was required and all data were anonymously collected in real-time, through an online server. On the 21^st^ day of practice, the rewards were removed (extinction) to ensure that the action sequences were more dependent on proprioceptive and kinesthetic, rather than on external, feedback. Analysis of the reward removal (extinction) is presented in the supplementary material. Other additional task components are also included in the supplementary materials.

#### Training engagement

Participants reliably committed to their regular training schedule, practicing consistently both sequences every day. Unexpectedly, OCD patients completed significantly more practices as compared with HV (*p* = 0.005) (Figure 2a). Descriptive statistics are as follows: HV: median number of practices, M*_P_ =* 122*, IQR (Interquartile range)* = 7; OCD: M*_P_ =* 130*, IQR* = 14. Note that as summary statistics we provide the median, and errors are reported as interquartile range unless otherwise stated, due to the non-Gaussian distribution of the datasets. When visually inspecting the daily training pattern, we observed that HV have a tendency to practice somewhat earlier than OCD. Circular statistics within each group demonstrated that HV practiced preferentially at a peak time of ∼15:00 (mean resultant length 0.47, p = 0.000497, Rayleigh test for the uniformity of a circular distribution of points; Figure 2b). For OCD participants, the preferred practice time had a mean direction at ∼18:00 (mean resultant length 0.58, p = 8.03 x 10−6, Rayleigh test; Figure 2c). There were, however, no significant differences between both samples (p = 0.19, Watson’s U2 test).

**Figure 2.**
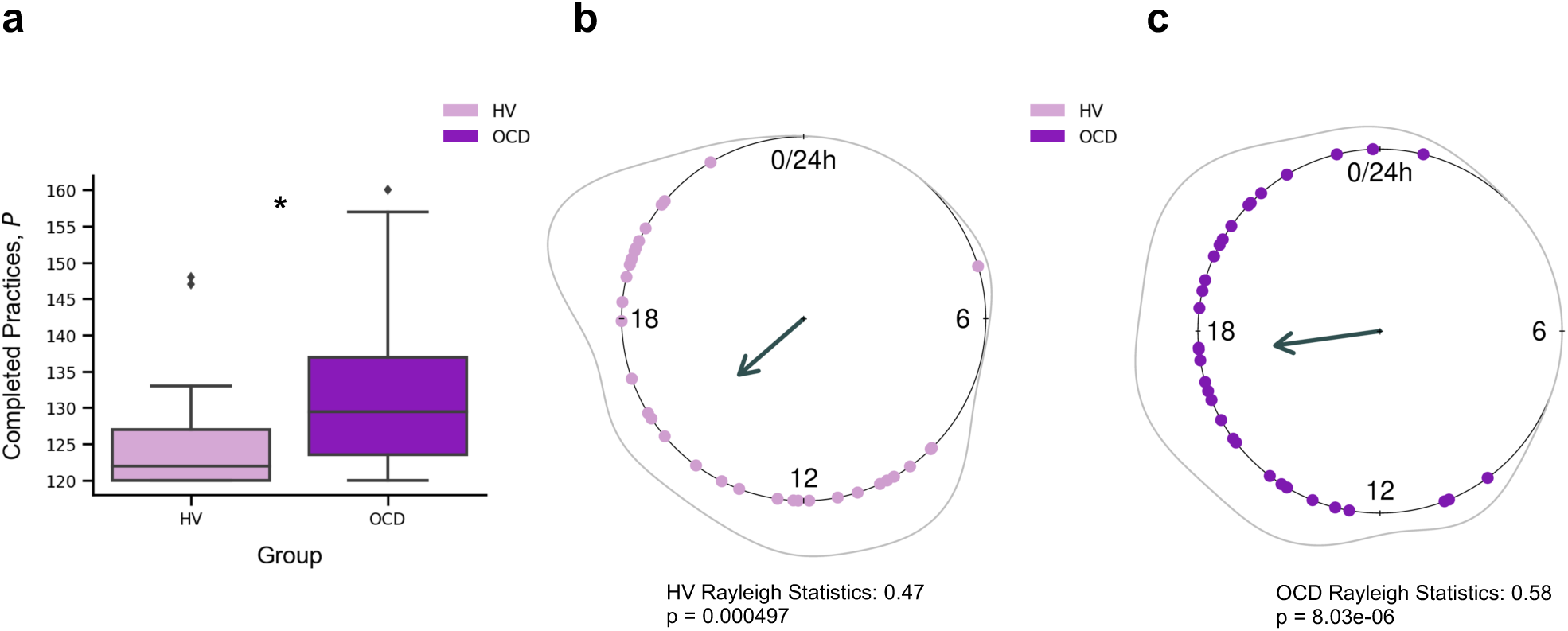
Training Engagement. **a)** Whole training overview. OCD patients engaged in significantly more training sessions than HV (* *p* = 0.005). The minimum required practices (*P*) were 120*P*. **b)** Daily training pattern for HV and **c)** Daily training pattern for OCD. Single dots on the unit circle denote the preferred practice times of individual participants within 0-24 hours, obtained from the mean resultant vector of individual practice hours data (Rayleigh statistics). Group-level statistics was conducted in each group separately using the Rayleigh test to assess the uniformity of a circular distribution of points. The graphic displays the length of the mean resultant vector in each distribution, and the associated p-value. Regarding between-group statistical analysis, see main text.

#### Learning

Learning was evaluated by the decrement in sequence duration throughout training. To follow the nomenclature of the motor control literature, we refer to sequence duration as movement time (*MT,* in seconds), which is defined as

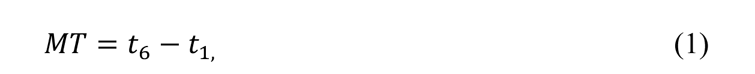

where *t*_6_and *t*_1_ are the time of the last (6^th^) and first key presses, respectively.

For each participant and sequence reward type (continuous and variable), we measured *MT* of a successful trial, as a function of the sequence trial number, *n,* across the whole training. Across trials, *MT* decreased exponentially (Figure 3a). The decrease in *MT* has been widely used to quantify learning in previous research (Crossman, 1959). A single exponential is viewed as the most statistically robust function to model such decrease (Heathcote et al., 2000). Accordingly, each participant’s learning profile was modeled as follows:

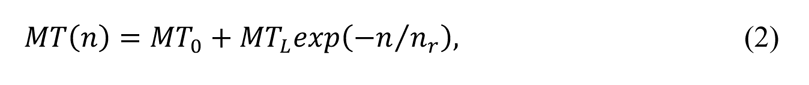

**Figure 3.**
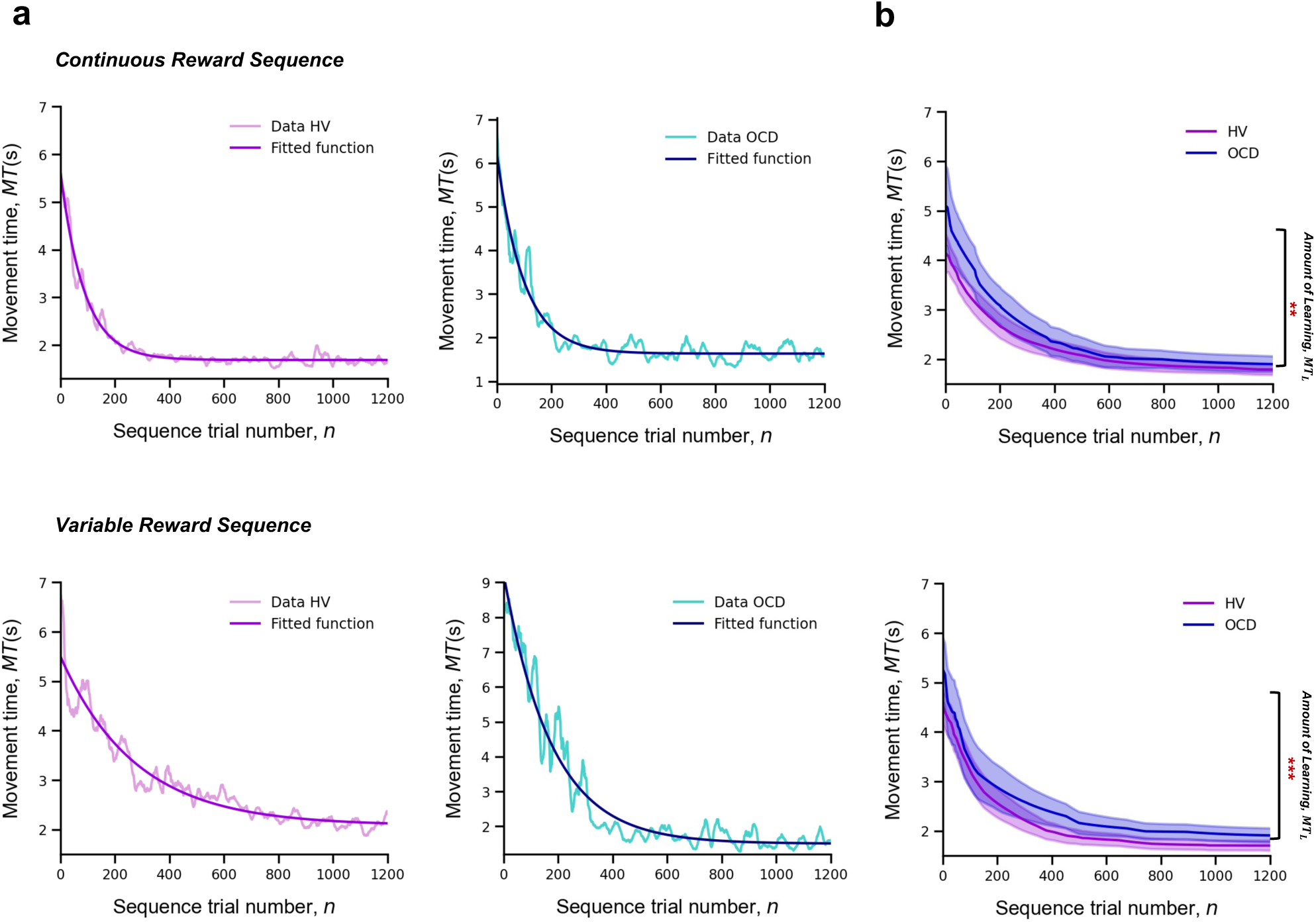
Learning. **Upper panel:** Model fitting procedure conducted for the continuous reward sequence. **Lower panel:** Model fitting procedure conducted for the variable reward sequence. **a)** Individual plots exemplifying the time-course *MT* (in seconds) as training progresses (lighter color) as well as the exponential decay fit modeling the learning profile of a single participant (darker color). Left panels depict data in a HV individual, right panels display data in a patient with OCD. **b)** Group comparison resulting from all individual exponential decays modeling the learning profile of each participant. A significant group difference was observed on the amount of learning, *MT*_*L*_, in both reward schedule conditions (continuous: *p* = 0.009; variable: *p* < 0.001). Solid lines: median (*M*); Transparent regions: median +/- 1.57 * interquartile range/sqrt(n); Purple: healthy volunteers (HV); Blue: patients with obsessive-compulsive disorder (OCD).

where *n* is the *learning rate* (measured in number of trials), which governs the rate of exponential decay. Parameter *MT*_0_is the movement time at *asymptote* (at the end of the training). Last, *MT*_*L*_ is the speed-up achieved over the course of the training (referred to as *amount of learning*) (Figure 3a). The larger the value of *MT*_*L*_, the bigger the decline in the movement time and thus the larger the improvement in motor learning.

The individual fitting approach we used has the advantage of handling the different number of trials executed by each participant by modeling their behavior to a consolidated maximum value of *n*, *n*_max_ = 1200. We used a moving average of 20 trials to mitigate any effect of outlier trials. This analysis was conducted separately for continuous and variable reward schedules.

To statistically assess between-group differences in learning behavior, we pooled the individual model parameters (*MT*_*L*_, *n*_r_, and *MT*_0_) , and conducted a Kruskal–Wallis H test to assess the effect of group (HV and OCD), reward type (continuous and variable) and their interaction on each parameter (Figure 3b).

There was a significant effect of group on the *amount of learning* parameter (*MT*_*L*_*, H* = 16.5, *p* < 0.001, but no reward (*p* = 0.06) or interaction effects (*p* = 0.34) (Figure 3c). Descriptive statistics: HV: M *MT*_*L*_= 3.1 s, *IQR* = 1.2 s and OCD: M *MT*_*L*_= 3.9 s, *IQR* = 2.3 s for the continuous reward sequence; HV: M *MT*_*L*_= 2.3 s, *IQR* = 1.2 s and OCD: M *MT*_*L*_= 3.6 s, *IQR* = 2.5 s for the variable reward sequence.

Regarding the *learning rate* (*n*_&_) parameter, we found no significant main effects of group (*p* = 0.79), reward (*p* = 0.47) or interaction effects (*p* = 0.46). Descriptive statistics: sequence trials needed to asymptote HV: M*n*_&_= 176, *IQR* = 99 and OCD: M*n*_&_= 200, *IQR* = 114 for the continuous reward sequence; HV: M*n*_&_= 182, *IQR* = 123 and OCD: M*n*_&_= 162, *IQR* = 141 for the variable reward sequence. These non-significant effects on the learning rate were further assessed with Bayes Factors (BF) for factorial designs (see Methods). This approach estimates the ratio between the full model, including main and interaction effects, and a restricted model that excludes a specific effect. The evidence for the lack of main effect of group was associated with a BF of 0.38, which is anecdotal evidence. We additionally obtained moderate evidence supporting the absence of a main effect of reward or a reward x group interaction (BF = 0.16 and 0.17 respectively).

In analyzing the asymptote parameter, *MT*_0_, we found no significant main or interaction effects (group effect: *p* = 0.17; reward effect: *p* = 0.65 and interaction effect: *p* = 0.64). Descriptive statistics are as follows: HV: M *MT*_0_= 1.7 s, *IQR* = 0.4 s and OCD: M *MT*_0_= 1.8 s, *IQR* = 0.5 s for the continuous reward sequence; HV: M *MT*_0_= 1.8 s, *IQR* = 0.5 s and OCD: M *MT*_0_= 1.8 s, *IQR* = 0.5 s for the variable reward sequence. BF analysis indicated anecdotal evidence against a main group effect (BF = 0.53). Meanwhile, there was moderate evidence suggesting neither reward nor reward x interaction factors significantly influence performance time (BF = 0.12 and 0.17, respectively).

The results indicate that OCD patients do not exhibit learning deficits. While they initially performed action sequences slower than the HV group, their learning rates ultimately matched those of HV. Both groups showed comparable movement durations at the asymptote. This suggests that, though OCD patients began at a lower baseline level of performance, they enhanced their motor learning to a degree that reached the same asymptotic performance as the controls.

#### Automaticity

To assess automaticity, the ability to perform actions with low-level cognitive engagement, we examined the decline over time in the consistency of inter-keystroke interval (IKI) patterns trial to trial. We mathematically defined IKI consistency as the sum of the absolute value of the time lapses between finger presses from one sequence to the previous one

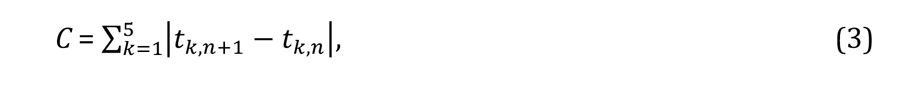

where *n* is the sequence trial number and *k* is the inter-keystroke response interval (Figure 4a). In other words, *C* quantifies how consistent/reproducible the press pattern is from trial to trial. The assumption here is that the more reproducible the sequences are *over time*, the more automatic the person’s motor performance becomes.

**Figure. 4.**
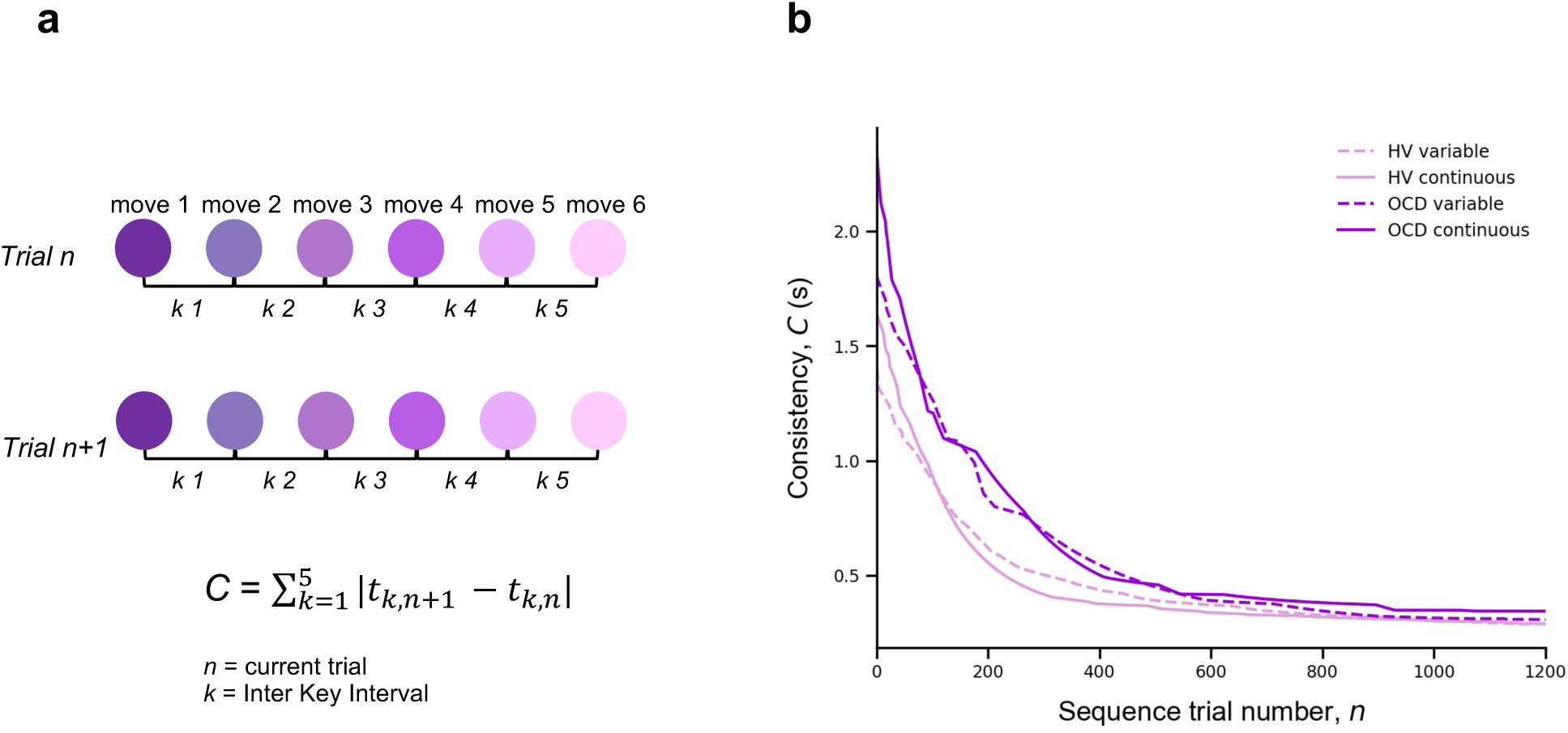
Automaticity. **a)** We mathematically defined trial-to-trial inter-keystroke-interval consistency (IKI consistency), denoted as *C* (in seconds), as the sum of the absolute values of the time lapses between finger presses across consecutive sequences. The variable *n* represents the sequence trial and *k* denotes the IKI. We evaluated automaticity by analyzing the decline in *C* over time, as it approached asymptotic levels. **b)** Group comparison resulting from all individual exponential decays modeling the automaticity profile (drop in *C*) of each participant. A significant group effect was found on the amount of automaticity gain, *C*_*L*_(Kruskal–Wallis *H* = 11.1, *p* < 0.001) and on the automaticity constant, *n*_)_ (Kruskal– Wallis H = 4.61, *p* < 0.03). Solid and dashed lines are median values (*M*). Light purple: healthy volunteers (HV); Dark purple: patients with obsessive-compulsive-disorder (OCD); Solid lines: continuous reward condition; Dashed lines: variable reward condition.

For each participant and sequence reward type (continuous and variable), automaticity was assessed based on the decrement in *C*, as a function of *n,* across the entire training period. Since *C* decreased in an exponential fashion, we fitted the *C* data with an exponential decay function (following the same reasoning and procedure as *MT*) to model the automaticity profile of each participant,

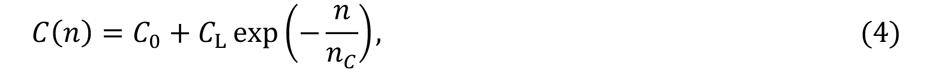

where *n*_)_ is the *automaticity rate* (measured in number of trials), *C*_*L*_is the sequence consistency at *asymptote* (by the end of the training) and *C*_*L*_is the change in automaticity over the course of the training (which we refer to *amount of automation gain*). The model fitting procedure was conducted separately for continuous and variable reward schedules.

A Kruskal–Wallis H test was then conducted to assess the effect of group (OCD and HV) and reward type (continuous and variable) on each parameter resulting from the individual exponential fits (*C*_*L*_, *n*_/_ and *C*_*L*_).

There was a significant effect of group on the *amount of automation gain* (*C*_*L*_: *H* = 11.1, *p* < 0.001) but no reward (*p* = 0.12) or interaction effects (*p* = 0.5) (Figure 4b). Descriptive statistics are as follows: HV: M*C*_*L*_= 1.4 s, *IQR* = 0.7 s and OCD: M*C*_*L*_= 1.9 s, *IQR* = 1.0 s for the continuous reward sequence; HV: M*C*_*L*_ = 1.1 s, *IQR* = 0.8 s and OCD: M*C*_*L*_= 1.5 s, *IQR* = 1.1 s for the variable reward sequence.

There was also a significant group effect on the *automaticity rate* (*n*_)_: *H* = 4.61*, p* < 0.03) but no reward (*p* = 0.42) or interaction (*p* = 0.12) effects. Descriptive statistics: sequence trials needed to asymptote HV: M*n*_)_= 142, *IQR* = 122 and OCD: M*n*_)_= 198, *IQR* = 162 for the continuous reward sequence; HV: M*n*_)_= 161, *IQR* = 104 and OCD: M*n*_)_= 191, *IQR* = 138 for the variable reward sequence.

At *asymptote,* no group (*p* = 0.1), reward (*p* = 0.9) or interaction (*p* = 0.45) effects were found. We found anecdotal evidence against a main group effect (BF = 0.65). In addition, there was moderate evidence in favor of no main effects of reward or interaction (BF = 0.12 and 0.18 respectively).

Of note is the median difference in consecutive sequences achieved at asymptote: HV: M*D*_0_ = 287 ms, *IQR* = 127 ms, OCD: *MD*_0_ =301 ms, *IQR* = 186 ms for the continuous reward sequence and HV: M*D*_0_ = 288 ms, *IQR* = 110 ms, OCD: M*D*_0_ = 300 ms, *IQR* = 114 ms for the variable reward sequence. These values of the *C* at asymptote are generally shorter than the normal reaction time for motor performance (Kosinski, 2008), reinforcing the idea that automaticity was reached by the end of the training.

In conclusion, compared to HV, patients took significantly longer to achieve a similar level of automaticity in both reward schedules. They began at a slower pace, exhibited more variability, and progressed to automaticity at a slower rate.

#### Sensitivity of sequence duration to reward

Our next goal was to investigate the sensitivity of performance improvements over time in our participant groups to changes in scores, whether they increased or decreased. To do this, we quantified the trial-by-trial behavioral changes in response to a decrement or increase in reward from the previous trial using the sequence duration (in ms), labeled as *MT* (movement time). Note that in our experimental design, *MT* was negatively correlated with the scores received. Following Pekny et al., 2015, we represented the change from trial *n* to *n + 1* in *MT* simply as:

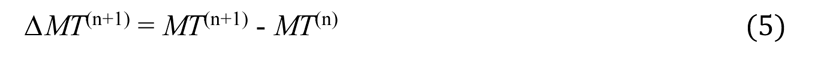

Reward (R) change at trial *n* was computed as:

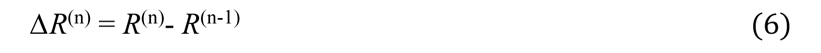

We next aimed to analyze separately *ΔMT* values that followed an increase in reward from trial *n − 1* to *n*, *ΔR+*, denoting a positive sign in *ΔR*; and those that followed a drop in reward, *ΔR−*, indicating a negative sign in *ΔR*. An issue arises with poor performance trials (those with a slower duration, or a large *MT*^(n)^). These could inherently result in a systematic link between *ΔR*− and smaller (negative) *ΔMT*^(n+1)^ values due to the statistical effect known as ’regression to the mean’. Essentially, a trial that is poorly performed, marked by a large *MT*^(n)^ is likely to be followed by a smaller *MT*^(n+1)^ just because extreme values tend to be followed by values closer to the mean. As training progresses and *MT* reduces overall, the potential for significant changes relative to reward increments or decrements may diminish. To account for and counteract this statistical artifact, we normalized the *ΔMT*^(n+1)^ index using the baseline *MT*^(n)^:

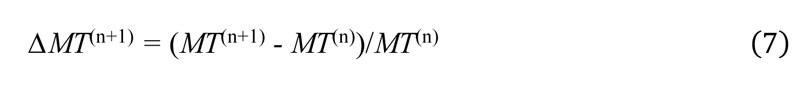

We used this normalized measure of *ΔMT*^(n+1)^ (adimensional) for further analyses. It reflects the behavioral change from trial *n* to *n+1* relative to the baseline performance on trial *n*. Following Pekny et al., 2015, we estimated for each participant the conditional probability distributions *p(ΔT|ΔR+)* and *p(ΔT|ΔR−)* (where *T* denotes a behavioral measure, *MT* in this section or *IKI* consistency in the next section) by fitting a Gaussian distribution to the histogram of each data sample (Figure S4). The standard deviation (*σ*) and the center *μ* of the resulting distributions were used for statistical analyses (Figure S4). Similar analyses were carried on a normalized version of index C (equation [3]), which already reflected changes between consecutive trials. See next section.

As a general result, we expected that healthy participants would introduce larger behavioral changes (more pronounced reduction in *MT,* more negative *ΔMT*) following a decrease in scores, as shown previously (Chen et al., 2017; van Mastrigt et al., 2020). Accordingly, we predicted that the *p(ΔT|ΔR−)* distribution would be centered at more negative values than p(ΔT|ΔR+), corresponding to greater speeding following negative reward changes. Given previous suggestions of enhanced sensitivity to negative feedback in patients with OCD (Apergis-Schoute et al., 2023, Becker et al., 2014; Kanen et al., 2019), we predicted that the OCD group, as compared to the control group, would demonstrate greater trial-to-trial changes in movement time and a more negative center of the *p*(*ΔT|ΔR*−) distribution. Additionally, we examined whether OCD participants would exhibit more irregular changes to ΔR− and ΔR+ values, as reflected in a larger spread of the *p*(*ΔT|ΔR*+) and p(*ΔT |ΔR* −) distributions, compared to the control group.

The conditional probability distributions were separately fitted to subsamples of the data across *continuous* reward practices, splitting the total number of correct sequences into four bins. This analysis allowed us to assess changes in reward sensitivity and behavioral changes across bins of sequences (bins 1-4 by partitioning the total number of sequences, from the whole training, into four). We focused the analysis on the continuous reward schedule for two reasons: 1) changes in scores on this schedule are more obvious to the participants and 2) a larger number of trials in each subsample were available to fit the Gaussian distributions, due to performance-related reward feedback being provided on all trials.

We observed that participants speeded up their sequence duration more (negative changes in trial-wise *MT*) following a drop in scores, as expected (Figure 5a). Conducting a 3-way ANOVA with reward change (increase, decrease) and bin (1:4, each bin denoting ∼110 sequences) as within-subject factors, and group as between-subject factor, we found a significant main effect of reward (*p* = 2.0 x 10^-16^). This outcome indicated that, in both groups, participants reduced *MT* differently as a function of the change in reward. There was also a significant main effect of bin (*p* = 5.06 x 10^-12^), such that participants speeded up their sequence performance over practices. The main effects are illustrated in Figure 5b. There was no significant main group effect (*p* = 0.2951), and omitting the group factor from the full model using a Bayes factor ANOVA analysis improved the model moderately (BF = 6.08, moderate evidence in support of the full model with the main group effect removed). Thus, both OCD and HV individuals introduced comparable changes in *MT* overall during training.

**Figure 5.**
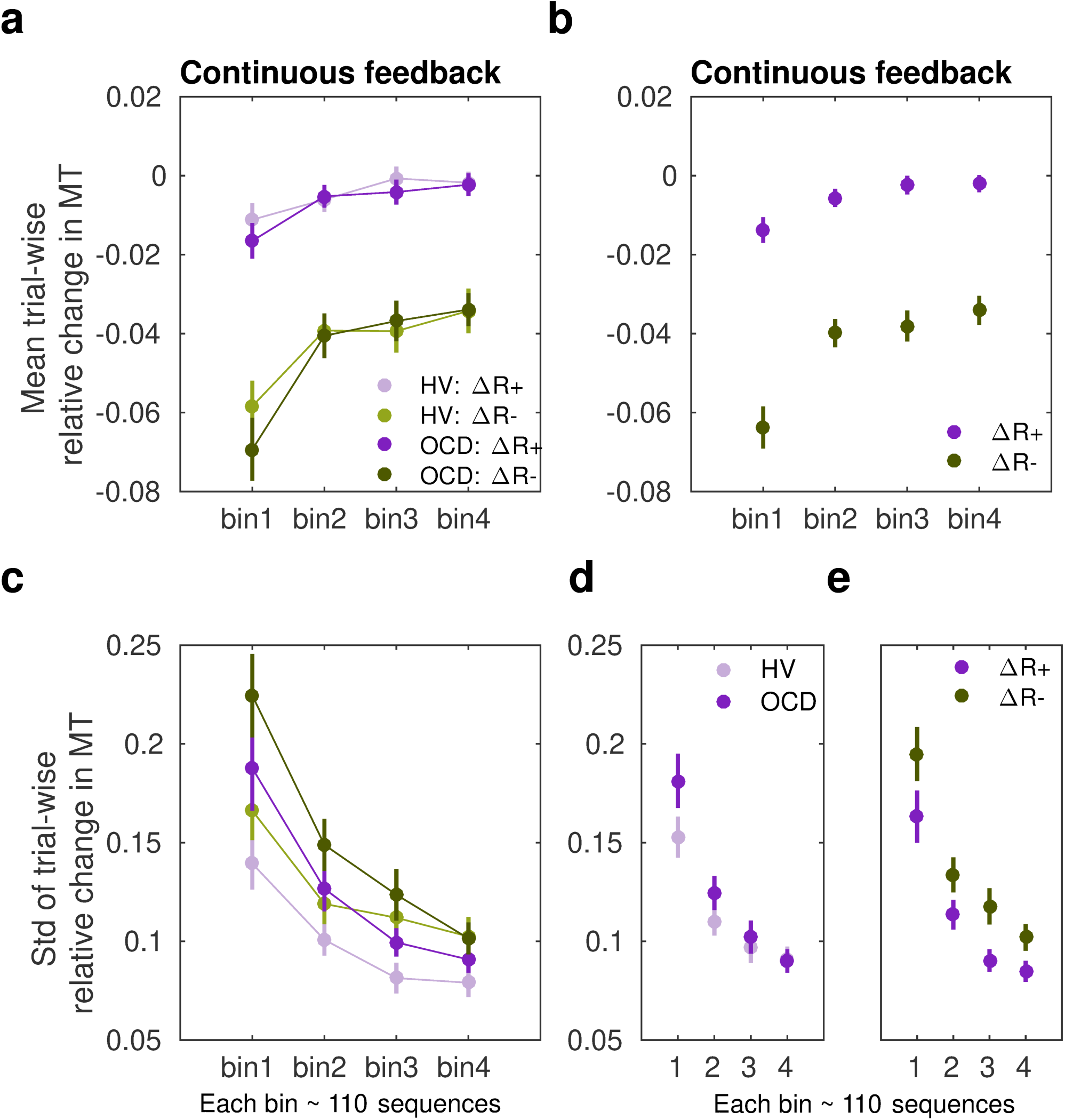
Sensitivity of movement time to changes in reward in the continuous reward schedule. **a)** Mean normalized change in movement time (*MT*, ms) from trial *n* to *n+1* following an increment (*ΔR+*, in purple) or decrement (*ΔR-*, in green) in scores at *n*. The change in movement time trial-to-trial was normalized with the baseline value on the initial trial *n*: *ΔMT^(n+1)^ = (MT^(n+1)^ - MT^(n)^*)/*MT^(n)^*. This relative change index is therefore adimensional. The dots represent mean *MT* changes (error bars denote SEM) in each bin of correctly performed sequences, after partitioning all correct sequences into four subsets, and separately for OCD (dark colors), and HV (light colors). **b)** Both groups of participants speeded up their sequence performance more following a drop in scores (main effect of reward, *p* = 2 x 10^-16^ ; 2 x 4: reward x bin ANOVA); yet this acceleration was reduced over the course of practiced sequences (main bin effect, *p* = 5.06 x 10^-12^). **c)** Same as a) but for the spread (std) of the *MT* change distribution (adimensional). **d-e)** Illustration of the main effect of group (**[d]** *p* = 9.93 x 10^-6^) and reward (**[e]** *p* = 4.13 x 10^-5^) on std. Each bin depicted in the plots (x-axis) contains around 110 correct sequences on average (further details in ***Supplementary Results: Sample size for the reward sensitivity analysis***).

In addition, there was a significant interaction between reward and bin in predicting the trial-to-trial changes in movement time (*p* = 0.0126). This outcome suggested that the relative improvement in *MT* over sequences depended on whether the reward increased or decreased from the previous trial. To explore this interaction effect further, we conducted a dependent-sample pairwise *t-*test on *MT*, after collapsing the data across groups. In each sequence bin, participants speeded up *MT* more following a drop in scores than following an increment, as expected (corrected *p_FDR_* = 2 x 10^-16^). On the other hand, assessing the effect of bins separately for each level of reward, we observed that the large sensitivity of normalized *MT* changes to reward decrements was attenuated from the first to the second bin of practice (corrected *p_FDR_* = 0.00034, significant attenuation for pairs 1-2; dependent-sample t-tests between consecutive pairs of bins). No further changes over practice bins were observed (*p* > 0.53, no change for pairs 2-3, 3-4). Similarly, the sensitivity of *MT* changes to reward increments—consistently smaller—did only change from bin 1 to bin 2 (*p_FDR_* = 0.01092; no significant changes for pairs 2-3 and 3-4, *p* > 0.31670).

Overall, these findings indicate that both OCD and HV participants exhibited an acceleration in sequence performance following a decrease in scores (main effect). Furthermore, the sensitivity to score decrements or increments was reduced as participants approached automaticity through repeated practice. Crucially, however, the increased sensitivity to reward decrements relative to increments persisted throughout the practice sessions in both groups.

Assessment of the std (*σ*) of the Gaussian distributions *p (ΔT |ΔR−)* and *p(ΔT |ΔR+)* in the continuous reward condition (Figure 5c) with a similar 3-way ANOVA revealed a significant main effect of group (*p* = 9.93 x 10^-6^). As shown in Figure 5d, the std (*σ*) of the distribution of trial-to-trial *MT* changes was smaller in HV than in OCD. In addition, we observed a significant change over bins of sequences in (*σ*), and independently of the group or reward factors (main effect of bin, *p* = 2 x 10^-16^). This outcome reflected that over practices, both groups introduced less variable changes in MT over the course of training in response to both reward increments and decrements, in line with improvements in skill learning (Wolpert et al., 2011). Reward also modulated *σ*, with *ΔR−* being associated with a more variable distribution of behavioral changes than *ΔR−* (main effect of reward, *p* = 4.13 x 10^-05^). No interaction effects were found (there was moderate to strong evidence that removing any of the possible interaction effects improved the model: BF ranged from 5.67 to 41.3). Control analyses demonstrated that the group, reward or bin effects were not confounded by differences in the size of the subsamples used for the Gaussian distribution fits (Supplementary Results).

#### Sensitivity of IKI consistency (C) to reward

To further explore the potential impact of reward changes on the previously reported group effects on automaticity, we quantified the trial-by-trial behavioral changes in IKI consistency (represented by *C*) in response to changes in reward scores relative to the previous trial. Note that a smaller C indicates a more reproducible IKI pattern trial to trial. As for *ΔMT*, we normalized the index *C (*termed *normC* to avoid confusion with the main analysis on *C)* with the baseline IKI values on the previous trial *n*

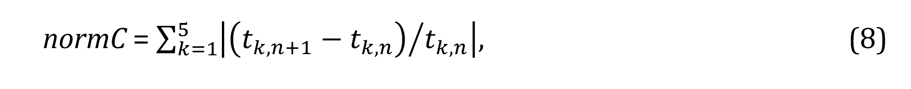

where *k* is the inter-keystroke response interval and *n* is the sequence trial number. During continuous reward practices, both patients and healthy controls exhibited an increased consistency of IKI patterns trial to trial across bins of correct sequences (decreased *normC*, equation [8], Figure 6a; main effect of bin on the center of the Gaussian distribution, *p* = 0.00607; 3-way ANOVA). Performance in OCD and HV, however, differed with regards to how reproducible their timing patterns were (main effect of group, *p* = 0.00454). The timing patterns were less consistent trial-to-trial in OCD, relative to HV (Figure 6b, left panel). Moreover, the IKI consistency improved more (smaller *normC*) following reward increments than after decrements, as shown in Figure 6b (right panel; main reward effect, *p* = 1.86 x 10^-06^). No significant interaction effects between factors were found (Bayes factor analysis demonstrated that when any of the interaction effects among factors was removed from the ANOVA design, there was moderate to strong evidence that the model improved: BF in range from 6.83 to 14.1). Accordingly, although OCD participants exhibited an attenuated IKI consistency in their performance relative to HV, the main effects of reward and bins of sequences were independent of the group.

**Figure 6.**
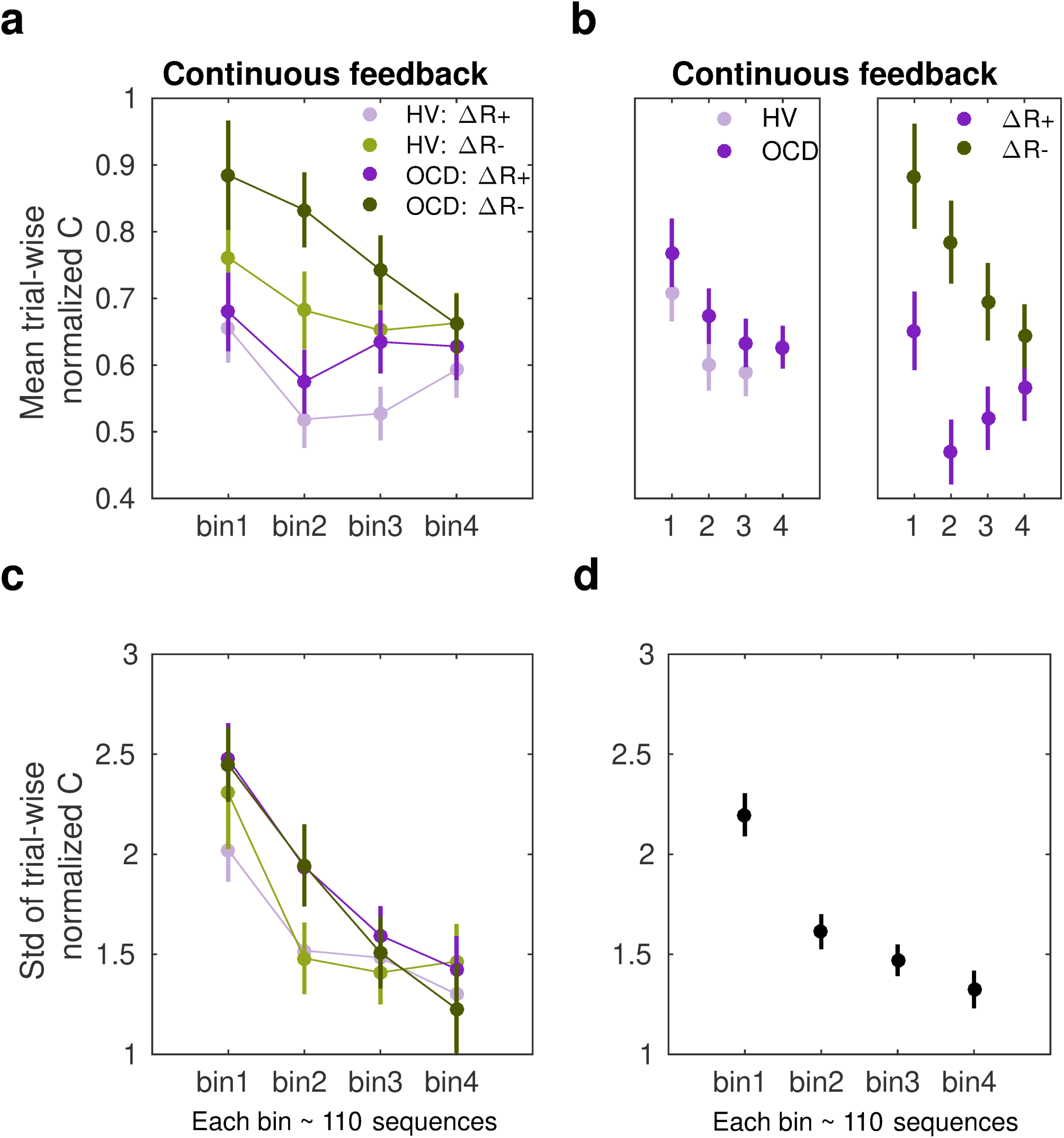
Sensitivity of normalized IKI consistency (norm*C*) to reward changes in the continuous schedule. **a)** The mean normalized change in trial-to-trial IKI consistency (*normC*, equation [8]; adimensional) across bins of correct sequences is shown, separately for each group (OCD: dark colors; HV: light colors) and for reward increments (*ΔR+*, purple) and decrements (*ΔR-*, green). The dots represent the mean value, while the vertical bars denote SEM. **b)** Illustration of the main effect of group (left panel; *p* = 0.00454) and type of reward change (right panel; *p* = 1.86 x 10^-06^). **c)** Same as a) but for the std of the distribution of IKI consistency changes, *normC*, adimensional. **d)** The panel displays the main effect of bin (*p* = 3.63 x 10^-14^) on the std. Black denotes the average (SEM) across reward and group levels. Each bin depicted in the plots (x-axis) contains 110 correct sequences on average (See ***Supplementary Results***: ***Sample size for the reward sensitivity analysis)*.**

Regarding the spread of the *p(ΔT |ΔR)* distributions, we found a significant main effect of bin factor (*p* = 3.63 x 10^-14^; Figure 6cd). This outcomes suggest that the σ of the Gaussian distribution for norm*C* values was reduced across bins of practiced sequences. There was only a trend for a significant main effect of group (*p* = 0.0653) and no main effect of reward (*p* = 0.1278). These non-significant effects were explored further using Bayes factors. This analysis provided anecdotal and moderate evidence that omitting either the group or reward effects was beneficial to the model (BF = 1.98 for removing group, BF = 3.20 for removing reward). We did not observe any interaction effect either (BF values increased moderately to strongly when any of the interaction effects among factors was removed from the ANOVA design: BF ranged from 4.89 to 43.3). The results highlight that over the course of training participants’ normalized IKI consistency values stabilized, and this effect was not observed to be modulated by group or reward factors. Similarly to the *MT* analyses, the sensitivity analyses of *normC* were not influenced by differences in the size of the subsamples used for the *ΔR+* and *ΔR-* Gaussian distribution fits (Supplementary Results).

#### Phase B: Tests of action-sequence preference and re-evaluation

Once the month-long app training was completed, participants attended a laboratory session to conduct additional behavioral tests aimed at assessing preference for familiar versus novel sequences (experiment 2 and 3) including a re-evaluation test to assess ability to adapt to environmental changes (experiment 3 only). Below we briefly describe these two experiments and report the results. See Methods and **Table 3** for a more detailed description of the tasks. Since these follow-up tests required observing additional stimuli *while* performing the action sequences, it was impractical to use participant’s individual iPhones to simultaneously present the task stimuli and be an interface to play the action sequences. We therefore used a ‘Makey-Makey’ device to connect the testing laptop (presenting the task stimuli) to four playdough keys arranged on a table (used as an interface for action sequence input). This device ensured precise key registration and timing. The playdough keys matched the size of those on the participants’ iPhones used for the one-month training. Participants practiced the action sequences in this new setup until they were comfortable. Hence, the transition to a non-mobile/laboratory context was conducted with great care. These tasks were conducted in a new context, which has been shown to promote re-engagement of the goal system (Bouton, 2021).

### Experiment 2

#### Preference for familiar versus novel action sequences

This experiment tests the hypothesis stated in the outline, that the trained action sequence gains intrinsic/rewarding properties or value. We used an *explicit preference task*, assessing participants’ preferences for familiar (hypothetically habitual) sequences over goal-seeking sequences. We assume that if the trained sequences have acquired rewarding properties (for example, anxiety relief, or the inherent gratification of skilled performance or routine), participants would express a greater preference to ‘play’ them, even when alternative easier sequences are offered (i.e., goal-seeking sequences).

After reporting which app sequence was their preferred, participants started the *explicit preference task*. On each trial, they were required to select and play 1 of 2 sequences. The 2 possible sequences were presented and identified using a corresponding image. Participants had to choose which one to play. There were 3 conditions, each comprising a specific sequence pair: 1) app preferred sequence *versus* app non-preferred sequence (*control condition*) 2) app preferred sequence *versus* any 6-move sequence (*experimental condition 1*); 3) app preferred sequence *versus* any 3-move sequence (*experimental condition 2*). The app preferred sequence was their preferred putative habitual sequence while the ‘any 6’ or ‘any 3’-move sequences were the goal-seeking sequences. These were considered less effortful for 2 reasons: 1) they could comprise any key press of participant’s choice, even repeated presses of the same key (6 or 3 times respectively), and 2) they allowed for variations in key combinations each time the ’any-sequence’ was input, rather than a fixed sequence on every trial. The conditions (15 trials each) were presented sequentially but counterbalanced among participants. See Methods and Figure 7a for further details.

**Figure 7.**
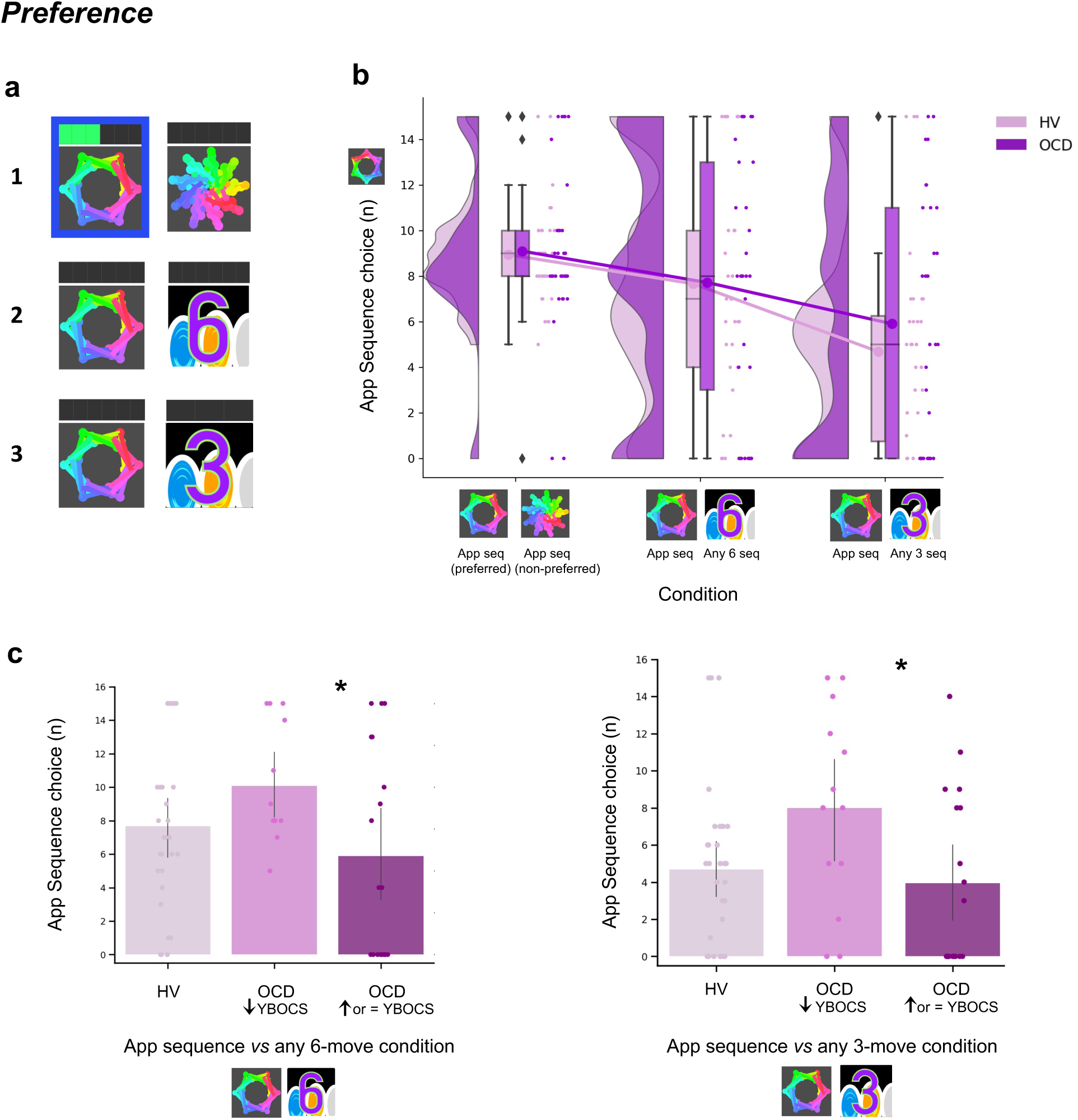
Preference for familiar versus novel action sequences. **a)** Explicit Preference Task. Participants had to choose and play one of two given sequences. Once choice was made, the image correspondent to the selected sequence was highlighted in blue. Participants then played the sequence. While playing it, the bar on top registered each move progressively lighting up in green. There were 3 conditions, each comprising a specific sequence pair: 1) app preferred sequence *versus* app non-preferred sequence (*control condition*) 2) app preferred sequence *versus* any 6-move sequence of participant’s choice (*experimental condition 1*); 3) app preferred sequence *versus* any 3-move sequence of participant’s choice (*experimental condition 2*). **b)** No evidence for enhanced preference for the app sequence in either HV nor OCD patients. In fact, when an easier and shorter sequence is pitted against the app familiar sequence (right raincloud plot), both groups significantly preferred it (Kruskal-Wallis main effect of Condition *H* = 23.2, *p* < 0.001). Left raincloud plot: control condition; Middle raincloud plot: experimental condition 1; Right raincloud plot: experimental condition 2. Y-axis depicts the number of app-sequence choices (15 choice trials maximum). Connected lines depict mean values. **(c)** Exploratory analysis of the preference task following up unexpected findings on the mobile-app effect on symptomatology: re-analysis of the data conducting a Dunn’s *post hoc* test splitting the OCD group into 2 subgroups based on their YBOCS change after the app training [14 patients with improved symptomatology (reduced YBOCS scores) and 18 patients who remained stable or felt worse (i.e. respectively, unchanged or increased YBOCS scores)]. Patients with reduced YBOCS scores after the app training had significantly higher preference to play the app sequence in both experimental conditions (left panel: *pFDR = 0.015*\*; right panel: *pFDR = 0.011*\*). The bar plots represent the sample mean and the vertical lines the confidence interval. Individual data points are included to show dispersion in the sample. Abbreviations: YBOCS = Yale-Brown obsessive-compulsive scale, HV = Healthy volunteers, OCD = patients with obsessive-compulsive disorder.

A Kruskal-Wallis H test indicated a main effect of Condition (*H* = 23.2, *p* < 0.001) but no Group (*p* = 0.36) or interaction effects (*p* = 0.72) (Figure 7b). Dunn’s *post hoc* pairwise comparisons revealed that *experimental condition 2 (app sequence versus any 3 sequence*) was significantly different from *control condition* (*p_FDR_ < 0.001)* and from *experimental condition 1 (app sequence versus any 6 sequence*) (*p_FDR_ = 0.006).* No differences were found between the latter two conditions (*p = 0.086).* Bayesian analysis further provided moderate evidence in support of the absence of main effects of group (BF = 0.129) and interaction (BF = 0.054). These results denote that both groups evaluate the trained app sequences as being equally attractive as the alternative novel-but-easier sequence when of the same length (Figure 7b, middle plot). However, when given the option to play an easier-but-shorter sequence (in *experimental condition 2*), both groups significantly preferred it over the app familiar sequence (Figure 7b, right plot). A positive correlation between COHS and the app sequence choice (*Pearson r* = 0.36, *p* = 0.005) showed that those participants with greater habitual tendencies had a greater propensity to prefer the trained app sequence under this condition.

Given the high variance of participants’ choices on this preference task, particularly in the experimental conditions, and the findings reported below related to the mobile-app performance effect on symptomatology, we further conducted an exploratory Dunn’s *post hoc* test splitting the OCD group into 2 subgroups based on their Yale-Brown Obsessive-Compulsive Scale (YBOCS) score changes after the app training: 14 patients with improved symptomatology (reduction in YBOCS scores) and 18 patients who remained stable or felt worse (i.e. respectively, same or increase in YBOCS scores). Patients with lowered YBOCS scores after the app training had significantly greater preference for the app trained sequence in both experimental conditions as compared to patients with same or increased YBOCS scores after the app training: *experimental condition 1* (*p*_FDR_= 0.015, Figure 7c, *left*) and *experimental condition 2* (*p*_FDR_= 0.011, Figure 7c, *right*). In addition to this subgrouping analysis, we conducted a correlation analysis between changes in YBOCS scores and patient preferences for the app-sequences. This helped us determine whether patients who experienced greater changes in YBOCS scores tended to prefer the learned sequences, and vice versa. We observed a positive correlation, meaning that the higher the symptom improvement after the month training, the greater the preference for the familiar/learned sequence. This is particularly the case for the experimental condition 2, when subjects are required to choose between the trained app sequence and any 3-move sequence (r_s_ = 0.35, *p* = 0.04). A trend was observed for the correlation between the YBOCS score change and the preference for the app-sequences in experimental condition 1 (r_s_ = 0.30, *p* = 0.09). In conclusion, most participants preferred to play shorter and easier alternative sequences, thus not showing a bias towards the trained/familiar app sequences. Contradicting our hypothesis, OCD patients followed the same behavioral pattern. However, some participants still preferred the app sequence, specifically those with greater habitual tendencies, including patients who improved their symptoms during the month training and considered the app training beneficial (see also below exploratory analyses of “Mobile-app performance effect on symptomatology”). Such preference presumably arose because some intrinsic value may have been attributed to the trained action sequence.

### Experiment 3

#### Re-evaluation of the learned action sequence

In Experiment 3, we employed a *<u>2-choice appetitive learning task</u>*. We modified the conditions by manipulating extrinsic feedback to assess participants’ capacity to adopt a different response choice, after re-evaluating their options. By providing more value to alternative action sequences (as opposed to the previously automatized ones), participants were thus encouraged to reassess their choices and respond appropriately. Of note, we did not use a conventional goal devaluation procedure here, as this could possibly have disrupted the behavioral control of the sequences and thus invalidated the test.

On each trial, participants were required to choose between two ‘chests’ based on their associated reward value. Each chest depicted an image identifying the sequence that needed to be completed to be opened. After choosing which chest they wanted, participants had to play the specific correct sequence to open it. Their task was to learn by trial and error which chest would give them more rewards (gems), which by the end of the experiment would be converted into real monetary reward. There was no penalty for incorrectly keyed sequences because behavior was assessed based on participants’ choice regardless of the sequence accuracy.

Four chest-pairs (conditions, 40 trials each) were tested (see Figure 8a and methods for detailed description of each condition): three conditions pitted the trained/familiar app sequence against alternative sequences of higher monetary outcomes (given by variable amount of reward that did not overlap [deterministic]). The fourth condition kept the monetary value equivalent for the two options (maintaining a probabilistic rather than deterministic contingency) but offered a significantly easier/shorter alternative sequence. This set up a comparison between the intrinsic value of the familiar sequence and a motor-wise less effortful sequence. The conditions were presented sequentially but counterbalanced among participants.

**Figure 8.**
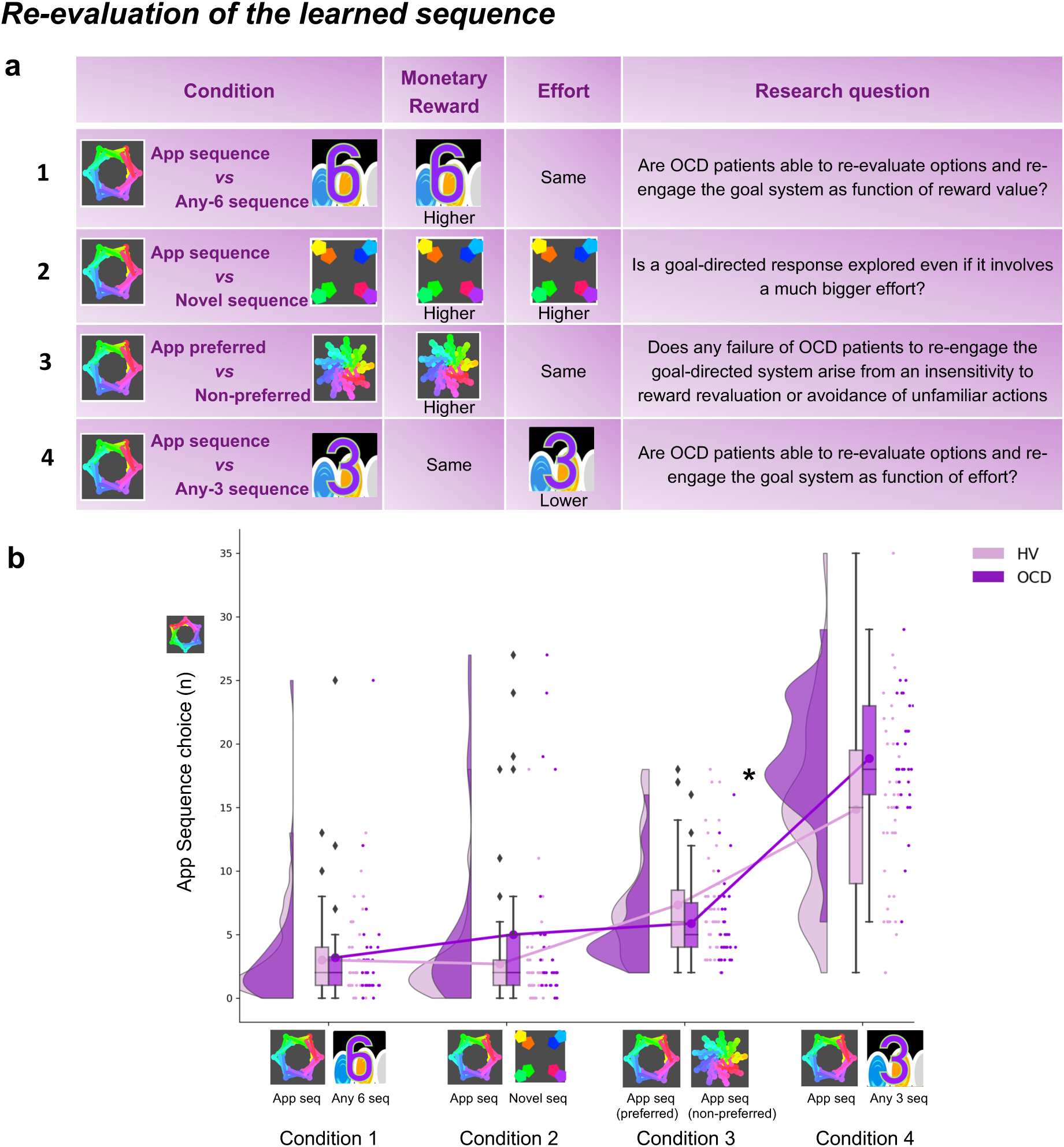
Re-evaluation procedure: 2-choice appetitive learning task. **a)** shows the task design. We tested 4 conditions, with chest-pairs corresponding to the following motor sequences: 1) app preferred sequence *versus* any 6-move sequence; 2) app preferred sequence *versus* novel (difficult) sequence; 3) app preferred sequence *versus* app non-preferred sequence; 4) app preferred sequence *versus* any 3-move sequence. The ‘any 6-move’ or ‘any 3-move’ sequences could comprise any key press of the participant’s choice and could be played by different key press combinations on each trial. The ‘novel sequence’ (in 2) was a 6-move sequence of similar complexity and difficulty as the app sequences, but only learned on the test day (therefore, not overtrained). In conditions 1, 2 and 3, the preferred app sequence was pitted against alternative sequences of higher monetary value. In condition 4, the intrinsic value of the preferred app sequence was pitted against a motor-wise less effortful sequence (i.e. a shorter/easier sequence). Each condition addressed specific research questions, which are detailed in the right column of the table. **b)** demonstrates the task performance per group and over the 4 conditions. Both groups were able to adjust to the new contingencies and choose the sequences the sequences associated to higher monetary reward. When re-evaluation involved an motor effort manipulation, OCD patients chose the app sequence significantly more than HV (* = *p* < 0.05) (condition 4). Y-axis depicts the number of app-sequence chests chosen (40 trials maximum) and connected lines depict mean values.

Both groups were highly sensitive to the re-evaluation procedure based on monetary feedback, choosing more often the non-app sequence, irrespective of the novelty of that sequence (Figure 8b, no group effects; *p* = 0.210 and BF = 0.742, anecdotal evidence supporting no main effect of group). However, when re-evaluation required motor effort (condition 4), participants were less inclined to choose the ’any 3’ alternative, which is the sequence demanding less motor effort (Kruskal-Wallis main effect of condition: *H* = 151.1 *p* < 0.001). Moreover, OCD patients significantly favored the trained app sequence over HV (*post hoc* group x condition 4 comparison: *p* = 0.04). In conclusion, following the month of training, both groups exhibited the ability to update their behavior based on monetary reevaluation. Yet, OCD patients more frequently selected the familiar sequence, even when a less effortful and shorter alternative was available.

### Mobile-app performance effect on symptomatology: exploratory analyses

In a debriefing questionnaire, participants were asked to give feedback about their app training experience and how it interfered with their routine: a) how stressful/relaxing the training was (rated on a scale from -100% highly stressful to 100% very relaxing); b) how much it impacted their life quality (Q) (rated on a scale from -100% maximum decrease to 100% maximum increase in life quality). Supplementary Table S1 and Figure S5 depicts participants’ qualitative and quantitative feedback. Of the 33 HV, 30 reported the app was neutral and did not impact their lives, neither positively nor negatively. The remaining 3 reported it as being a positive experience, with an improvement in their life quality (rating their life quality increase as 10%, 15% and 60%). Of the 32 patients assessed, 14 unexpectedly showed improvement (*I*) in their OCD symptoms during the month as measured by the YBOCS difference, in percentage terms, pre-post training (*I*= 20 ± 9%), 5 felt worse (*I*= -19 ± 9%) and 13 remained stable during the month (all errors are standard deviations). Of the 14 who felt better, 10 directly related their OCD improvement to the app training (life quality increase: *Q*= 43 ± 24%). Nobody rated the app negatively. Of note, the symptom improvement was positively correlated with patients’ habitual tendencies reported in the Creature of Habits questionnaire, particularly with the routine subscale (Pearson *r* = 0.45, *p* = 0.01) (Figure 9a, left). A three-way ANOVA test showed that patients who reported less obsessions and compulsions after the month training were the ones with more pronounced habit routines (Group effect: F = 13.7, *p* < 0.001, Figure 9a, right). A strong positive correlation was also found between the OCD improvement reported subjectively as direct consequence of the app training and the OCI scores and reported habit tendencies (Pearson *r* = 0.8, *p* = 0.008; Pearson *r* = 0.77, *p* < 0.01, respectively) (Figure 9b): i.e., patients who considered the app somewhat beneficial were the ones with higher compulsivity scores and higher habitual tendencies. In HV, participants who also had greater tendency for automatic behaviors, regarded the app as more relaxing (Pearson *r* = 0.44, *p* < 0.01). However, such correlation between the self-reported relaxation measure attributed to the app and the COHS automaticity subscale was not observed in OCD (*p* = 0.1). Finally, patients’ symptom improvement did not correlate with how relaxing they considered the app training (*p* = 0.1) nor with the number of total practices performed during the month training period (*p* = 0.2).

**Figure 9.**
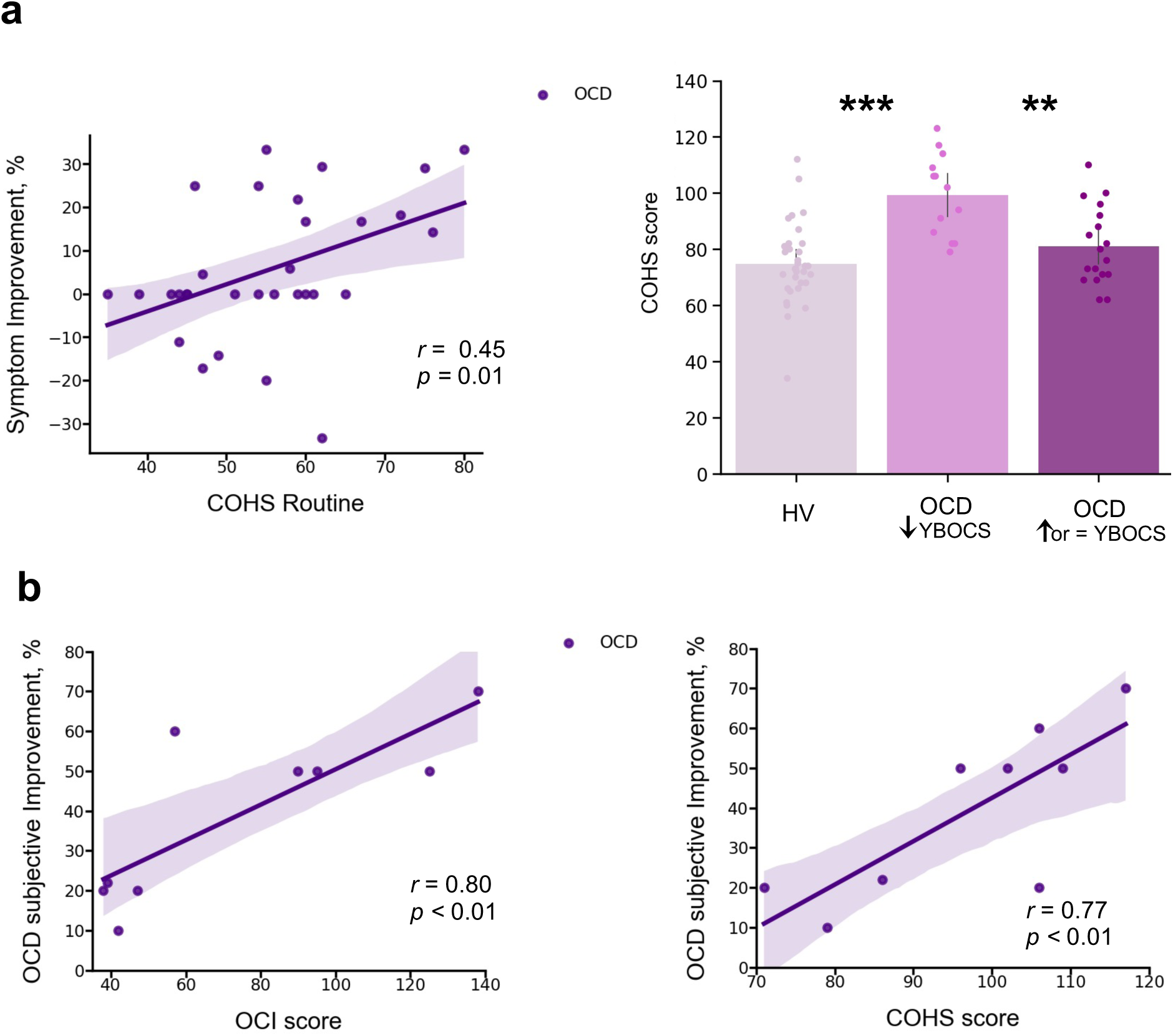
Mobile-app effect on symptomatology. **a)** *Left:* positive correlation between patients’ routine tendencies reported in the Creature of Habits (COHS) questionnaire and the symptom improvement (Pearson *r* = 0.45, *p* = 0.01). Symptom improvement was measured by the difference in YBOCS scale before and after app training. *Right*: Patients with greater improvement in their symptoms after the one month app training had greater habitual tendencies as compared to HV (*p* < 0.001) and to patients who did not improve post-app training (*p* = 0.002). The bar plot represents the sample means and the vertical lines the confidence interval. Individual data points are included to show dispersion in the sample. **b)** OCD patients who related their symptom improvement directly to the app training were the ones with higher compulsivity scores on the OCI (Pearson *r* = 0.8, *p* = 0.008) (*left*) and higher habitual tendencies on the COHS (Pearson *r* = 0.77, *p* < 0.01) (*right*). Note that b) has one missing patient because he did not complete the OCI and COHS scales. **Abbreviations**: OCI = Obsessive-Compulsive Inventory, COHS = Creature of Habits Scale, YBOCS = Yale-Brown obsessive-compulsive scale, HV = Healthy volunteers, OCD = patients with obsessive-compulsive disorder.

We also checked whether the preferred app sequence, chosen by participants at the beginning of Phase B, was consistently the one that had yielded more reward during the app training (i.e. the continuously rewarded sequence). We found no evidence for this case: 54.5% of HV and 29% of the OCD sample considered the continuous sequence to be their preferred one, a non-statistically significant difference. This result suggests that participants’ preference may not solely be linked to programmed reward. Other factors, such as the aesthetic appeal of, or ease of performing specific combinations of finger movements, may also influence overall preference.

### Other self-reported symptoms

In addition to the Creature of Habit findings, of the remaining self-reported questionnaires assessed (see Methods), OCD patients also reported enhanced intolerance of uncertainty, elevated motivation to avoid aversive outcomes and higher perfectionism, worries and perceived stress, as compared to healthy controls (see Table 1 for statistical results and Figure 10 in the Methods section for overall summary).

**Figure 10.**
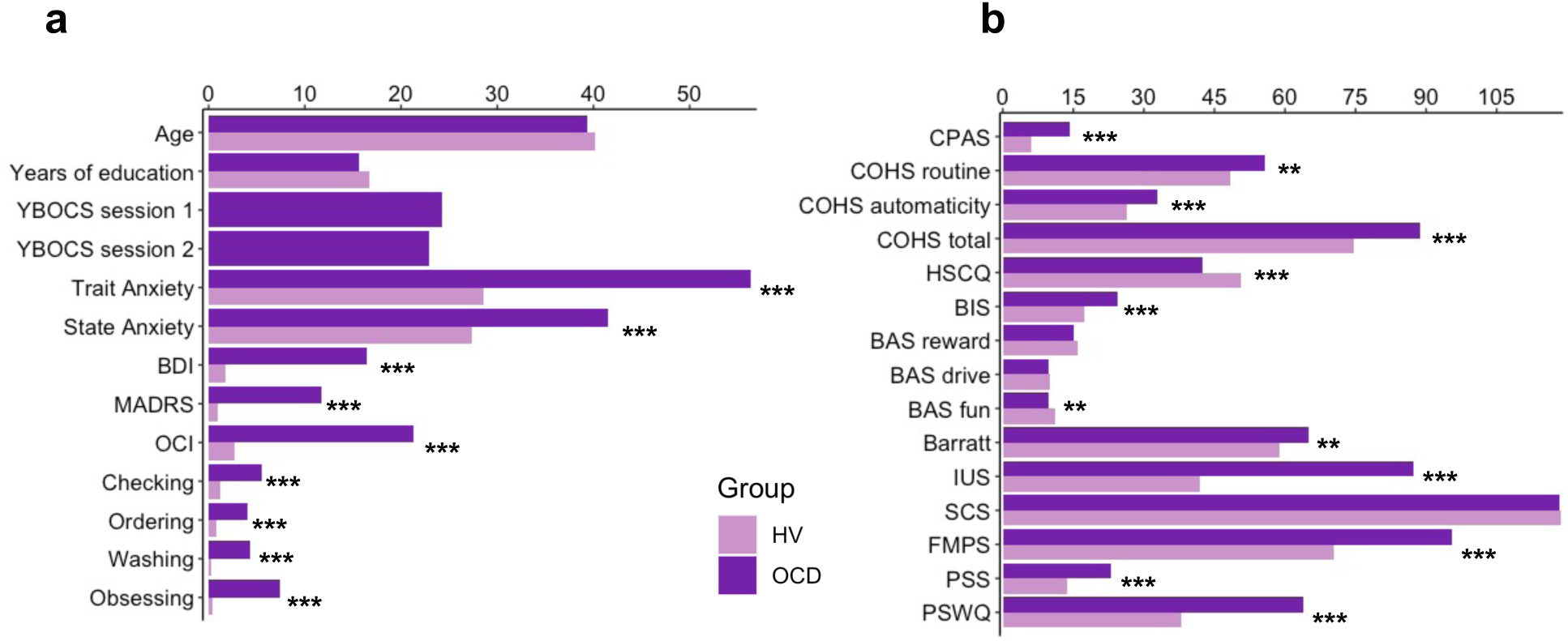
**a)** Participants’ demographics and clinical characteristics. **b)** Between group results from the self-reported questionnaires. **Abbreviations**: HV, Healthy Volunteers; OCD, Patients with Obsessive-Compulsive Disorder; Y-BOCS, Yale-Brown obsessive-compulsive scale; MADRS, Montgomery-Asberg Depression Rating Scale; STAI, The State-Trait Anxiety Inventory; BDI, Beck Depression Inventory; OCI, Obsessive-Compulsive Inventory; CPAS, Compulsive Personality Assessment Scale; COHS, Creature of Habit Scale; HSCQ, Habitual Self Control Questionnaire; BIS, Behavioral Inhibition System; BAS, Behavioral Activation System; Barratt, Barratt Impulsiveness Scale; IUS, Intolerance of Uncertainty Scale; SCS, Self-Control Scale; FMPS, Frost Multidimensional Perfectionism Scale; PSS, Perceived Stress Scale; PSWQ, Penn State Worry Questionnaire. ** = *p* < 0.01, *** = *p* <0.001.

**Table 1.** Self-reported measures on various scales measuring impulsiveness, compulsiveness, habitual tendencies, self-control, behavioral inhibition and activation, intolerance of uncertainty, perfectionism, stress and the trait of worry.

|  | HV<br>(n= 33) | OCD<br>(n= 32) | t | Statistics<br>df | p |
| --- | --- | --- | --- | --- | --- |
| CPAS | 5.9 (4.0) | 14.2 (5.0) | -7.37 | 62 | <0.001*** |
| COHS routine | 48.4 (9.4) | 55.7 (11.1) | -2.79 | 62 | 0.01* |
| COHS automaticity | 26.3 (8.2) | 32.9 (8.5) | -3.15 | 62 | <0.001*** |
| COHS total | 74.8 (14.4) | 88.7 (16.7) | -3.56 | 62 | <0.001*** |
| HSCQ | 50.7 (7.3) | 42.5 (8.5) | 4.17 | 62 | <0.001*** |
| BIS | 17.5 (3.5) | 24.4 (2.7) | -8.81 | 61 | <0.001*** |
| BAS reward responsibility | 15.9 (2.2) | 15.1 (2.5) | 1.25 | 61 | 0.22 |
| BAS drive | 10.0 (2.4) | 9.6 (2.6) | 0.66 | 61 | 0.51 |
| BAS fun seeking | 11.1 (1.9) | 9.7 (2.4) | 2.60 | 61 | 0.01* |
| Barratt total | 58.8 (8.4) | 65.0 (10.1) | -2.68 | 61 | 0.01* |
| Barratt attentional | 14.6 (4.1) | 19.8 (4.7) | -4.74 | 61 | <0.001*** |
| Barratt motor | 21.2 (2.6) | 21.4 (3.2) | -0.23 | 61 | 0.82 |
| Barratt non-planning | 23.7 (3.3) | 24.6 (4.5) | -0.96 | 61 | 0.34 |
| IUS | 41.9 (10.0) | 87.3 (20.2) | -11.23 | 61 | <0.001*** |
| SCS | 118.5 (21.4) | 118.3 (17.2) | 0.04 | 62 | 0.97 |
| FMPS | 70.3 (21.0) | 95.4 (21.4) | -4.73 | 62 | <0.001*** |
| PSS | 13.7 (4.7) | 22.9 (5.1) | -7.51 | 62 | <0.001*** |
| PSWQ | 37.9 (11.7) | 64.0 (11.0) | -9.20 | 62 | <0.001*** |

**Abbreviations:** HV, Healthy Volunteers; OCD, Patients with Obsessive-Compulsive Disorder; CPAS, Compulsive Personality Assessment Scale; COHS, Creature of Habit Scale; HSCQ, Habitual Self Control Questionnaire; BIS, Behavioral Inhibition System; BAS, Behavioral Activation System; Barratt, Barratt Impulsiveness Scale; IUS, Intolerance of Uncertainty Scale; SCS, Self-Control Scale; FMPS, Frost Multidimensional Perfectionism Scale; PSS, Perceived Stress Scale; PSWQ, Penn State Worry Questionnaire. Standard deviations are in parentheses: mean (std). One patient and one healthy control missed a few questionnaires. \* = $p < 0.05$ level, \*\*\* = $p < 0.001$ level (2-tailed).

## Discussion

This study investigated the roles of habits, their automaticity, and potential adjustments to environmental changes underlying compulsive OCD symptoms. We specifically focused on the habitual component of the associative dual-process model of behavior as applied to OCD, and described in the

Introduction. Using a self-report questionnaire (Ersche et al., 2017), we observed heightened subjective habitual tendencies in OCD patients across both the ’routine’ and ’automaticity’ domains, in comparison to controls.

Leveraging a novel smartphone tool, we real-time monitored the acquisition of two putative ’procedural’ habits (6-element action sequences) in OCD patients and healthy participants over 30 days in their daily environments. Our analyses revealed heightened engagement with the app training among OCD patients; they enjoyed and practiced the sequences more than healthy participants without any explicit directive to do so. Initially, these patients performed the sequences more slowly and irregularly, yet they eventually achieved the same asymptotic level of automaticity and exhibited comparable ’chunking’ (Smith and Graybiel, 2016) to controls. There were no discernible procedural learning deficits in patients, although their progression to automaticity was significantly slower than in healthy participants.

In a subsequent testing phase in a novel context, both groups adeptly transferred both trained action sequences to corresponding discriminative stimuli (visual icons). Furthermore, both cohorts were sensitive to re-evaluation when it pertained to monetary reward, demonstrating their ability to adapt behavior when facing environmental changes. However, when re-evaluation involved physical effort, OCD patients did not demonstrate the same adaptability and instead displayed a distinct inclination toward the already trained/familiar action sequence, presumably due to its inherent value. This effect was more pronounced in patients with higher habitual inclinations and compulsivity scores. Exploratory analysis revealed that patients with pronounced habitual inclinations and compulsivity scores were more likely to choose the familiar sequence. Moreover, when faced with a choice between the familiar and a new, less effort-demanding sequence, the OCD group leaned toward the former, likely due to its inherent value. These insights align with the theory of goal-direction/habit imbalance in OCD (Gillan et al., 2016), underscoring the dominance of habits in particular settings where they might hold intrinsic value. This inherent value could hypothetically be associated with symptom alleviation. Corroborating this, post-training feedback and a measured difference in the Y-BOCS scale preand post-training suggest many patients found the app therapeutically beneficial.

### Implications for the dual associative theory of habitual and goal-directed control

Rapid execution, invariant response topography, action chunking and low cognitive load, have all been considered essential criteria for the definition of habits (Balleine and Dezfouli, 2019; Haith and Krakauer, 2018). We have successfully achieved all of these elements with our app using the criteria of extensive training and context stability, both previously shown to be essential to enhance formation and strengthening of habits (Haith and Krakauer, 2018; Verplanken and Wood, 2006). *Context stability* was provided by the tactile, visual and auditory stimulation associated with the phone itself, which establishes a strong and similar context for all participants, regardless of their concurrent circumstances. *Overtraining* has been one of the most important criteria for habit development, and used by many as an operational definition on how to form a habit (Dickinson et al., 1995; Haith and Krakauer, 2018; Tricomi et al., 2009) (for a review see Balleine and O’Doherty, 2010), despite current controversies raised by de Wit et al., 2018 on its use as an objective test of habits. A recent study has demonstrated though that even short overtraining (1 day) is effective at producing habitual behavior in participants high in affective stress (Pool et al., 2022), confirming previous suggestions for the key role of anxiety and stress on the behavioral expression of habits (Dias-Ferreira et al., 2009; Hartogsveld et al., 2020; Schwabe and Wolf, 2009). Here we have trained a clinical population with moderately high baseline levels of stress and anxiety, with training sessions of a higher order of magnitude than in previous studies (de Wit et al., 2018, 2018; Gera et al., 2022). By all accounts our overtraining is valid: to our knowledge the longest overtraining in human studies achieved so far. Al participants attained automaticity, exhibiting similar and stable asymptotic performance, both in terms of speed and the invariance in the kinematics of the motor movement.

We succeeded in achieving automaticity – which at a neural level is known to reliably engage the brain’s habitual circuitry (Ashby et al., 2010; Bassett et al., 2015; Graybiel and Grafton, 2015; Lehericy et al., 2005) – and fulfilled three of the four criteria for the definition of habits according to Balleine and Dezfouli 2019 (Balleine and Dezfouli, 2019) (rapid execution, invariant topography and chunked action sequences). We were not, however, able to test the fourth criterion of resistance to devaluation. Therefore, we are unable to firmly conclude that the action sequences are habits rather than, for example, goal-directed skills. According to a very recent study, also employing an app to study habitual behavior, the criterion of devaluation resistance was shown to apply to a 3-element sequence with less training (Gera et al., 2022). Thus, overtraining of our 6-element sequence might also have achieved behavioral autonomy from the goal in addition to behavioral automaticity. While we did not employ the conventional goal devaluation test, it is possible that some experts may interpret our follow-up Experiment 3 (the re-evaluation test) as a measure of Balleine and Dezfouli’s (2018) fourth criteria, which defines habits as “*insensitive to changes in their relationship to their individual consequences and the value of those consequences*”. Consequently, they may conclude that the app-trained sequences exhibited some features of goal-directed behavior. While this interpretation holds merit, the logical conclusion is that the app-trained sequences encompass both habitual and goaldirected qualities. This aligns with contemporary perspectives on skilled/habitual sequences (Du and Haith, 2023).

Regardless of whether the trained action sequences are labeled as procedural habits or goal-directed motor skills, one must question why OCD patients preferred familiar sequences in specific situations, even when it seemed counterproductive (e.g., in the effort condition). This observation leads to the hypothesis that motivation for action sequences might include other factors besides explicit goals, such as monetary rewards. The apparent (intrinsic) therapeutic value of performing these sequences further blurs the attribution of a singular goal such as monetary reward to human action sequences. One implication of this analysis may be to consider that behavior in general is ‘goal-directed’ but may vary in the balance of control by external and internal goals. This perspective aligns with motor control theories that classify the successful completion of a motor action, in the spatio-temporal sense, as ’goal-related’. Hence, underlying any action sequence is possibly a hierarchy of objectives, ranging from overt rewards like money to intrinsic relief from an endogenous state (e.g. anxiety or boredom). In light of these insights, the dual associative process framework of behavioral control might be better understood in terms of the relative importance of extrinsic versus intrinsic outcomes. Another possible formulation is that habits, which depend initially on cached or historically acquired rewarding action values may not necessarily lose current value, but instead acquire alternative sources of value (Hommel and Wiers, 2017; Kruglanski and Szumowska, 2020; O’Doherty, 2014).

### Implications for understanding OCD symptoms

We observed a slower and more irregular performance in patients with OCD as compared to healthy participants at the beginning of training. This was expected given previous reports of visuospatial and fine-motor skill difficulties in patients with OCD (Bloch et al., 2011). However, despite this initial slowness, no procedural learning deficits were found in our patient sample. This finding is inconsistent with other implicit learning deficits previously reported in OCD using the serial reaction time (SRT) paradigm (Deckersbach et al., 2002; Joel et al., 2005; Kathmann et al., 2005; Rauch et al., 2001, 1997). Nevertheless, this result aligns with recent studies demonstrating successful learning both in patients with OCD (Soref et al., 2018) and healthy individuals with subclinical OCD symptoms (Barzilay et al., 2022) when instructions are given explicitly and participants intentionally search for the underlying sequence structure. In fact, our task does not tap into memory processes as strongly as SRT tasks because we explicitly demonstrate the sequence to participants before they begin their 30-day training, which likely decreases demands on procedural learning.

Quantifying trial-to-trial behavioral changes in response to a decrease or increase in reward suggested that the slower progression towards automaticity observed in OCD patients might be related to their more inconsistent response to changes in feedback scores compared to healthy participants. The adjustments that OCD participants made to sequence duration after a score change were more variable (with a larger σ) than those made by healthy individuals. Additionally, the normalized consistency index was higher for the OCD group than for HV, indicating more fluctuating changes from trial to trial in inter-keystroke intervals. Despite these group differences, we observed that in both samples the consistency of IKI patterns improved after reward increments. This observation contrasts with the more pronounced MT acceleration in both groups when faced with negative reward changes.

A heightened sensitivity to negative feedback within the motor domain has been documented in the general population, influencing initial motor improvements, while an increase in reward primarily boosts motor retention (Abe et al., 2011; Galea et al., 2015; Pekny et al., 2015; van Mastrigt et al., 2020). OCD individuals have also been shown to have an amplified sensitivity to negative feedback (Becker et al., 2014). Our findings indicate that decreased feedback scores affect sequence duration and IKI consistency in distinct ways. Specifically, reduced score feedback hampered automatization (reducing the IKI consistency, increasing *normC)*, even though it generally had a positive effect on movement speed. This heightened responsiveness of *MT* (the rewarded variable) to decreased feedback scores is consistent with recent studies (Abe et al., 2011; Galea et al., 2015; Pekny et al., 2015; van Mastrigt et al., 2020). Our results, however, do not support differences in OCD and healthy individuals with regards to sensitivity to negative score changes, unlike previous work highlighting increased response switching after negative feedback, hyperactive monitoring systems, and amplified prediction errors in OCD (Hauser et al., 2017; Marzuki et al., 2021). One possible interpretation of these divergent results relates to the type of feedback. Previous work in OCD employed explicit negative feedback. In contrast, our participants received positive reward feedback, which increased or decreased trial-by-trial in the continuous reward schedule. The implicit nature of a reduced positive score, which is fundamentally different from overt negative feedback, might not elicit the same heightened sensitivity in OCD patients. They may primarily respond to explicit indications of failure or error, as opposed to subtle reductions in positive feedback. Another possibility is that the salient responses to negative feedback in OCD could be specific to the early stages of learning and may not persist after training for more than 1 hour or on subsequent days. Follow-up work will address these questions explicitly.

Considering the hypothetically greater tendency in OCD to form habitual/automatic actions described earlier (Gillan et al., 2014; Voon et al., 2015), we predicted that OCD patients would attain automaticity faster than healthy controls. This was not the case. In fact, the opposite was found. Since this was the first study to our knowledge assessing action sequence automatization in OCD, our contrary findings may confirm recent suggestions that previous studies were tapping into goal-directed behavior rather than habitual control per se (Gillan et al., 2015b; Vaghi et al., 2018; Zwosta et al., 2018) and may therefore have inferred enhanced habit formation in OCD as a defaulting consequence of impaired goal-directed responding. On the other hand, we are describing here two potential sources of evidence in favor of enhanced habit formation in OCD. First, OCD patients show a bias towards the previously trained, apparently disadvantageous, action sequences. In terms of the discussion above, this could possibly be reinterpreted as a narrowing of goals in OCD (Robbins et al., 2019) underlying compulsive behavior, in favor of its intrinsic outcomes. Secondly, OCD patients self-reported greater habitual tendencies in both the ‘routine’ and ‘automaticity’ subscales. Previous studies have reported that subjective habitual tendencies are associated with compulsive traits (Ersche et al., 2019; Wuensch et al., 2022) and act, in addition to cognitive inflexibility, as a predictor of subclinical OCD symptomatology in healthy populations (Ramakrishnan et al., 2022). There is an apparent discrepancy between self-reported ‘automaticity’ and the objective measure of automaticity we provided. This may result from a possible mis-labelling of this factor in the Creature of Habit questionnaire, where many of the relevant items indicate automatic S-R elicitation by situational triggering stimuli rather than motor topographic features of the behavior (e.g. ‘*when walking past a plate of sweets or biscuits, I can’t resist taking one*’).

Finally, we also expected that OCD patients would show a greater resistance than controls in adjusting their behavior when the extrinsic relative value of the trained familiar sequences is diminished, in the re-evaluation procedure. Our findings show that this is partially the case, depending on the type of reward considered. Although we showed that all participants, including OCD patients, were apparently goal-directed in terms of gaining money this was not so clear when goal re-evaluation involved the physical effort expended. In this latter manipulation, participants were less goal-oriented and OCD patients preferred to perform the longer, familiar, to the shorter, novel sequence, thus exhibiting significantly greater habitual tendencies, as compared to controls. Such group differences may be driven hypothetically by the intrinsic value associated with the automatic sequences.

### Possible beneficial effect of action sequence training on OCD symptoms as habit reversal therapy

OCD patients engaged significantly more with the motor sequencing app and enjoyed it more than healthy volunteers. Additionally, those patients were more prone to routine habits (COHS), had higher OCI scores and additionally showed a preference for familiar sequences, possibly by attributing to them intrinsic value, found use of the app beneficial, and exhibited symptomatic improvement based on the YBOCS scale. One hypothesis for the therapeutic potential of this motor sequencing training is that the trained action sequences may disrupt OCD compulsions either via ’distraction’ or habit ’replacement’ by engaging the same neural ‘habit circuitry’. This habit ’replacement’ hypothesis is in line with successful interventions in Tourette Syndrome (Hwang et al., 2012), Tic disorders and Trichotillomania (Morris et al., 2013).

### Limitations

As mentioned above, we were unable to employ the often-mooted ‘gold standard’ criterion of resistance to devaluation because it would have invalidated the subsequent tests. This meant that we were unable to conclusively define the trained action sequences as habitual according to the full set of Balleine and Dezfouli’s (2018) criteria, although they satisfied other important criteria such as automatic execution, invariant response topography and action chunking and low cognitive load. Nevertheless, the utility of the devaluation criterion has been questioned especially when applied to human studies of habit learning because devaluation can be difficult to achieve given that human behavior has multiple goals, some of which may be implicit, and thus difficult to control experimentally, as well as being subject to great individual variation. In fact recent analyses of habitual behavior have not employed devaluation or revaluation as a criterion (Du and Haith, 2023). That study ascertains habits using different criteria and provides supporting evidence for trained action sequences being understood as skills, with both goal-directed and habitual components.

Although we found a significant preference for the trained action sequence in OCD patients in the condition where it was pitted against a simpler and shorter motor sequence, as compared to the monetary discounting condition, the reason for this difference is not immediately obvious. However, it may have arisen because of the nature of the contingencies inherent in these choice tests. Specifically, the ‘monetary discounting’ condition involved a simple deterministic choice between the two alternatives, which should readily be resolved in favor of the option associated with the greater, non-overlapping, range of rewards provided (e.g. 1-7 versus 8-15 gems). In contrast, in the ‘effort discounting’ condition, the reward ranges for the two options were equivalent (e.g. 1-7 gems), which raised uncertainty concerning which of the chosen sequences was optimal. The probabilistic constraint over this choice may therefore account for the greater sensitivity of the task in highlighting preference in OCD, given the greater susceptibility of such patients to uncertainty (Pushkarskaya et al., 2015).

Finally, some of the conclusions relating to the effects of OCD diagnosis on sequence preference without feedback were based only on a *post hoc* exploratory analysis. Specifically, only those patients with higher compulsivity (OCI) and Creature Of Habit (COHS) scores exhibited this preference, therefore consistent with the hypothesis described above of the importance of intrinsic value of the habitual sequence to the development of compulsions. Evidence of this intrinsic value was provided by the greater engagement with, and therapeutic findings for, the app training in these patients. However, the latter effect needs to be confirmed in a registered clinical trial in a controlled manner, which is ongoing.

## Conclusion

We employed a battery of behavioral tasks designed to investigate two key hypotheses of the goal/habit imbalance theory of compulsion, specifically pertaining to enhanced habit formation and automaticity and impaired goal re-evaluation in individuals with OCD. Our findings did not support greater habit formation nor heightened automaticity in patients with OCD. Moreover, evidence for patients’ ability to adapt behavior when facing environmental changes was mixed. In certain contexts, OCD patients were able to behaviorally re-adjust (e.g. when reward is monetary) but in others (e.g. when involving motor effort) patients demonstrated a distinct augmented inclination to perform their trained/familiar action sequences, attributing higher intrinsic value to them. Interestingly, this preference was more pronounced in patients with higher compulsivity and habitual tendencies, who engaged significantly more with the motor habit-training app, reporting symptom relief after the experiment. This suggests a promising avenue for investigating the therapeutic potential of this application as a tool for habit reversal in the context of OCD.

## Materials and Methods

### Participants

We recruited 33 OCD patients and 34 healthy individuals, matched for age, gender, IQ and years of education. Two participants (1 HV and 1 OCD) were excluded because they did not perform the minimum required training (i.e. 2 daily practices for a period of 30 days). Therefore, a total of 32 OCD patients (19 females) and 33 healthy participants (19 females) were included in the analysis. Most participants were right-handed (left-handed: 4 OCD and 6 HV). Participants’ demographics and clinical characteristics are presented in Table 2 and Figure 10.

**Table 2.**
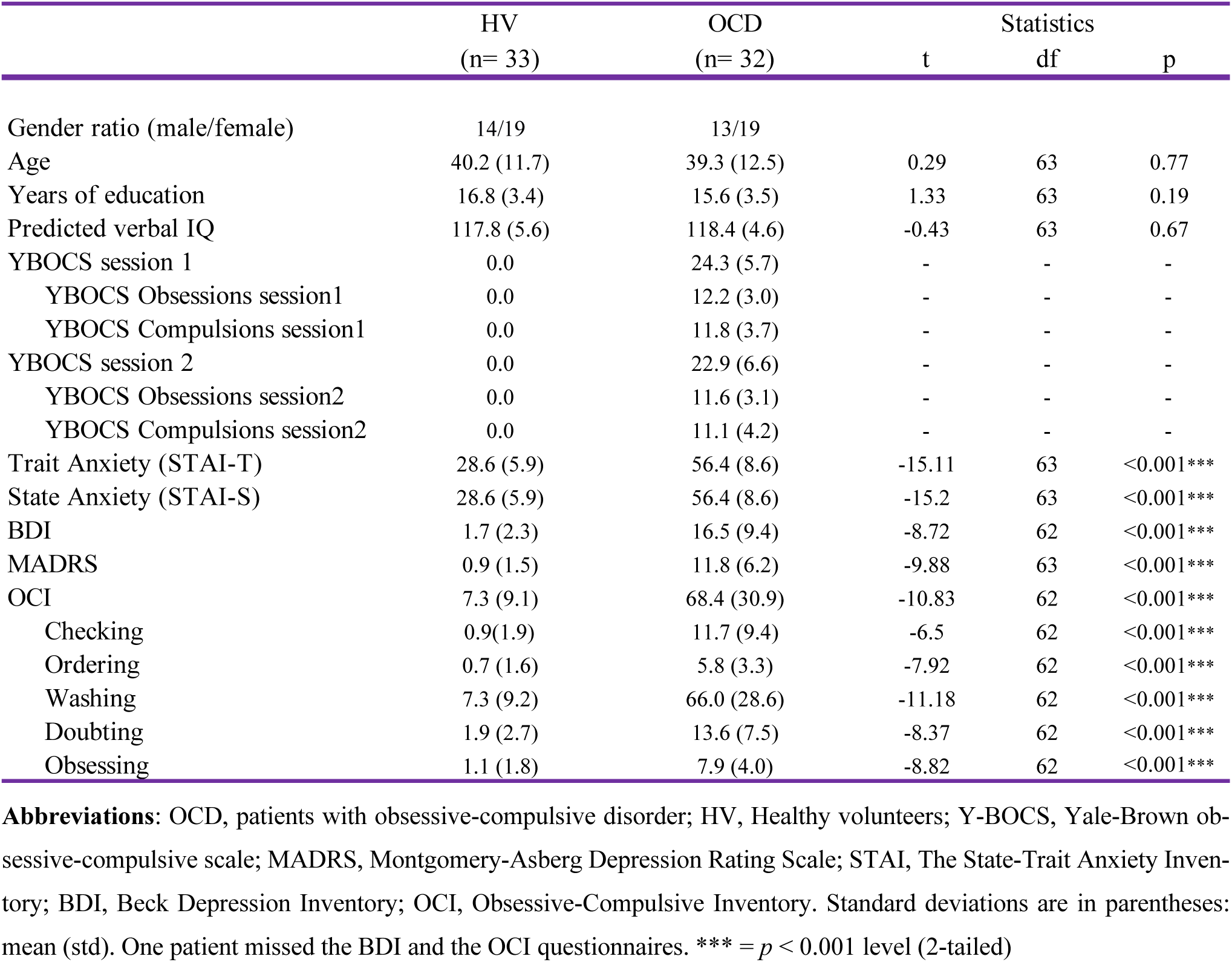
Demographic and clinical characteristics of OCD patients and matched healthy controls.

**Table 3.**
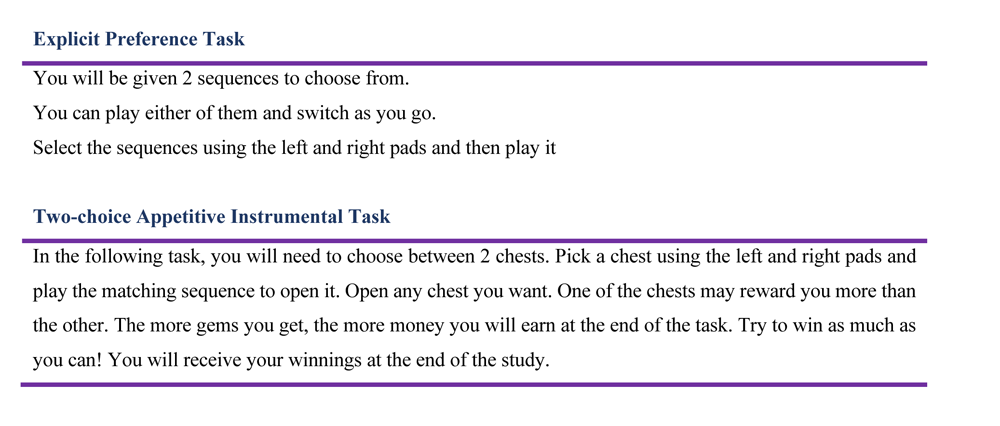
Follow up task instructions.

Healthy individuals were recruited from the community, were all in good health, unmedicated and had no history of neurological or psychiatric conditions. Patients with OCD were recruited through an approved advertisement on the OCD action website (www.ocdaction.org.uk) and local support groups and via clinicians in East Anglia. All patients were screened by a qualified psychiatrist of our team, using the Mini International Neuropsychiatric Inventory (MINI) to confirm the OCD diagnosis and the absence of any comorbid psychiatric conditions. Patients with hoarding symptoms were excluded. Our patient sample comprised 6 unmedicated patients, 20 taking selective serotonin reuptake inhibitors (SSRIs) and 6 on a combined therapy (SSRIs + antipsychotic). OCD symptom severity and characteristics were measured using the Y-BOCS scale (Goodman, 1989), mood status was assessed using the Montgomery-Asberg Depression Rating Scale (MADRS) (Montgomery and Asberg, 1979) and Beck Depression Inventory (BDI) (Beck et al., 1961), anxiety levels were evaluated using the State-Trait Anxiety Inventory (STAI) (Spielberger et al., 1983), and verbal IQ was quantified using the National Adult Reading Test (NART) (Nelson and Willison, 1982). All patients included suffered from OCD and scored > 16 on the Y-BOCS, indicating at least moderate severity. They were also free from any additional axis-I disorders. General exclusion criteria for both groups were substance dependence, current depression indexed by scores exceeding 16 on the MADRS, serious neurological or medical illnesses or head injury. All participants completed additional self-report questionnaires measuring:

a. impulsiveness: Barratt Impulsiveness Scale (Barratt, 1994)
b. compulsiveness: Obsessive Compulsive Inventory (Foa et al., 1998) and Compulsive Personality Assessment Scale (Fineberg et al., 2007)
c. habitual tendencies: Creature of Habit Scale (Ersche et al., 2017)
d. self-control: Habitual Self-Control Questionnaire (Schroder et al., 2013) and Self-Control Scale (Tangney et al., 2004)
e. behavioral inhibition and activation: BIS/BAS Scale (Carver and White, 1994)
f. intolerance of uncertainty (Buhr and Dugas, 2002)
g. perfectionism: Frost Multidimensional Perfectionism Scale (Frost and Marten, 1990)
h. stress: Perceived Stress Scale (Cohen et al., 1983)
i. trait of worry: Penn State Worry Questionnaire (Meyer et al., 1990).

All participants gave written informed consent prior to participation, in accordance with the Declaration of Helsinki, and were financially compensated for their participation. This study was approved by the East of England Cambridge South Research Ethics Committee (16/EE/0465).

### Phase B: Tests of action-sequence preference and re-evaluation

#### Experiment 2: explicit preference task

Participants observed, on each trial, 2 sequences identified by a corresponding image, and were asked to choose which one they wanted to play. Once choice was made, the image correspondent to the selected sequence was highlighted in blue. Participants then played the sequence. The task included 3 conditions (15 trials each). Each condition comprised a specific sequence pair: 2 experimental conditions pairing the app preferred sequence (putative procedural habit) with a goal-seeking sequence and 1 control condition pairing both app sequences trained at home. The conditions were as follows: 1) app preferred sequence *versus* app non-preferred sequence (*control condition*) 2) app preferred sequence *versus* any 6-move sequence (*experimental condition 1*); 3) app preferred sequence *versus* any 3-move sequence (*experimental condition 2*). The app preferred sequence was the putative habitual sequence and the ‘any 6’ or ‘any 3’-move sequences were the goal-seeking sequences because they are supposedly easier: they could comprise any key press of participant’s choice (for example the same single key press repeatedly 6 or 3 times respectively) and they could have same or different key press combinations every time the ‘any-sequence’ needed to be input. The conditions (15 trials each) were presented sequentially but counterbalanced among participants. Figure 7a for illustration of the task.

### Experiment 3: two-choice appetitive learning task

On each trial, participants were presented with two ‘chests’, each containing an image identifying the sequence that needed to be completed to be able to open the chest. Participants had to choose which chest to open and play the correct sequence to open it. Their task was to learn by trial and error which chest would give them more rewards ‘gems’, which by the end of the experiment would be converted into real monetary reward. If mistakes were made inputting the sequences, participants could simply repeat the moves until they were correct, without any penalty. Behavior was assessed based on participants’ choice, regardless of the accuracy of the sequence. The task included 4 conditions (40 trials each), with chest-pairs correspondent to the following motor sequences (see also figure 8 for illustration of each condition):

- *condition 1*: app preferred sequence *versus* any 6-move sequence
- *condition 2*: app preferred sequence *versus* a novel (difficult) sequence
- *condition 3*: app preferred sequence *versus* app non-preferred sequence
- *condition 4*: app preferred sequence *versus* any 3-move sequence

As in the preference task described above, the ‘any 6-move’ or ‘any 3-move’ sequences could comprise any key press of participant’s choice (for example the same single key press repeatedly 6 or 3 times respectively) and could be played by different key press combinations on each trial. The novel sequence (in condition 2) was a 6-move sequence of similar complexity and difficulty as the app sequences, but only learned on the day, before starting this task (therefore, not overtrained). The training of this novel sequence comprised 40 trials only: a number sufficient to learn the sequence without overtraining. Initially lighted keys guided the learning (similarly to the app training). After the initial 5 trials, the lighted cues were removed and participants were required to input the previously well learned correct 6-move sequence. When an error occurred, the correct input key(s) lighted up on the following trial (a few milliseconds before participants’ made key presses), to remind participants of the correct sequence and help them consolidate learning of the novel sequence. In conditions 1, 2 and 3, higher monetary outcomes were given to the alternative sequences. To remove the uncertainty confound commonly linked to probabilistic tasks, conditions 1, 2 and 3 followed a deterministic nature: in all trials, the choice for the preferred app sequence was rewarded with smaller monetary outcomes (sampled from a random distribution between 1-7 gems) whereas the alternative option always provided higher monetary outcomes (sampled from a random distribution between 8-15 gems). Therefore, variable amount of reward that did not overlap was given (deterministic). Condition 4, on the other hand, kept the monetary value equivalent for the two options (maintaining a probabilistic rather than deterministic contingency) but offered a significantly easier/shorter alternative sequence. This set up a comparison between the intrinsic value of the familiar sequence and a motor-wise less effortful sequence. To prevent excessive memory load, which could introduce potential confounds, conditions were presented sequentially rather than intermixed, but the order was counterbalanced among participants.

### Statistical analyses

Participant’s characteristics and self-reported questionnaires were analyzed with ξ_2_ and independent t-tests respectively. The Motor Sequencing App automatically uploaded the data to a cloud-based database. This task enabled us to compare patients with OCD and healthy volunteers in the following measures: training engagement (which included as primary output measures of the *total number of practices completed* and *app engagement* as defined as the number of sequences attempted, including both correct and incorrect sequences); procedural learning, automaticity development, sensitivity to reward (see definitions and description of data analyses in results section) and training effects on symptomatology as measured by the YBOCS difference pre-post training. The Phase B experiments enabled further investigation of preference and re-evaluation strategies. The primary outcome was the number of choices.

Between-group analyses were conducted using Kruskal-Wallis H tests when the normality assumption was violated. Parametric factorial analyses were carried out with analyses of variance (ANOVA). Our alpha level of significance was 0.05. On the descriptive statistics, main values are represented as median, and errors are reported as interquartile range unless otherwise stated, due to the non-Gaussian distribution of the datasets. When conducting several tests related to the same hypothesis, or when running several post-hoc tests following factorial effects, we controlled the FDR at level q = 0.05. Significant values after FDR control are denoted by *p*_FDR_. Analysis were performed using Python version 3.7.6 and JASP version 0.14.1.0.

In the case of non-significant effects in the factorial analyses, we assessed the evidence in favor or against the full factorial model relative to the reduced model with Bayes Factors (BF: ratio BFfull/BFrestricted) using the bayesFactor toolbox (https://github.com/klabhub/bayesFactor) in MATLAB®. This toolbox implements tests that are based on multivariate generalizations of Cauchy priors on standardized effects(Rouder et al., 2012). As recommended by Rouder and colleagues (2012), we defined the restricted models as the full factorial model without one specific main or interaction effect. The ratio BFfull/BFrestricted represents the ratio between the probability of the data being observed under the full model and the probability of the same data under the restricted model. BF values were interpreted following(Andraszewicz et al., 2015). The relationship between primary outcomes and clinical measures was calculated using a Pearson correlation.

The diurnal patterns of app use (Figure 2b and 2c) were assessed in each group using circular statistics (Mardia, 1975), with the “circular” package in R (R version 4.3.1; 2023-06-16). This provided the group-level mean vector length and direction. To assess on the group level whether the daily practice data were uniformly distributed or, alternatively, oriented towards a specific time, we used a Rayleigh-test (Landler et al., 2021; Mardia, 1975). We adapted code from (Galvez-Pol et al., 2022). To test for differences between two circular distributions (OCD, HV), we followed the recommendations of Landler et al., 2021 and employed the high-powered Watson’s U^2^ test, a non-parametric rank-based test (function watson.two.test in R).

## Data Availability

The source data for all figures and analyses are provided with this paper. They are available in the Open Science Framework, in the following link: https://osf.io/9xrdz/

## Acknowledgments

This research was funded by the Wellcome Trust: a Sir Henry Postdoctoral Research Fellowship (Grant 204727/Z/16/Z) to PB and a Wellcome Trust Senior Investigator Award (Grant 104631/Z/14/Z) to TWR. For the purpose of open access, the author has applied a CC BY public copyright licence to any Author Accepted paper version arising from this submission. MB was supported by MHRUK and Angharad-Dodds Bursaries. AAM was supported as a research assistant funded by the aforementioned Wellcome Trust grant TWR. We thank all participants for their contributions to this study. We would also like to acknowledge Dr Sharon Morein-Zamir for fruitful brainstorm discussions on the tasks design.

