## Supplemental File for "Action-sequence learning, habits and automaticity in obsessive-compulsive disorder"

**Supplementary Results**

### Motor Sequencing App: additional task components and analysis

The Motor Sequencing App comprised additional components and analysis, which we describe and report below respectively.

Once the minimum daily practice sessions were completed, we additionally asked participants to further conduct a short retention speed test, to assess that day's performance, and a switching practice session. In the 5-trial retention speed session, participants were instructed to repeatedly tap a sequence as rapidly as possible while making as few errors as possible. The 10 trial-switch session required switching between the two sequences in a pseudo-random order. The sequence to be played was cued by the respective associated picture. Speed and switch tests never received reward feedback (only the practice sessions). Finally, participants were asked to rate daily, on a percentage scale, the following two questions: (1) *How much did you enjoy playing this sequence?* and (2) *How confident are you that you know this sequence by heart?* This sequence of events (practice, speed, switch sessions and ratings) happened every day.

### Confidence and enjoyment ratings

We conducted a mixed-design repeated measures ANOVA to investigate potential group differences (OCD versus HV) on ratings of confidence (C) and enjoyment (E) over time (4 weeks of app practice). We observed a main effect of time on confidence [$F(3, 177) = 92.45$, $p < 0.001$, $\eta_p^2=0.19$] but no Group ($p = 0.61$) or interaction ($p = 0.95$) effects (Figure S1, a). This means that both groups significantly increased their confidence on their sequence knowledge over the course of the training. A Greenhouse-Geisser (ε) correction was applied given that sphericity was violated. Descriptive statistics are as follows: HV: $\overline{C}_{week1} = 64.03\% \pm 16.09$, $\overline{C}_{week2} = 74.90\% \pm 17.52$, $\overline{C}_{week3} = 80.14\% \pm 14.58$, $\overline{C}_{week4} = 85.60\% \pm 12.90$ and OCD: $\overline{C}_{week1} = 61.66\% \pm 20.62$, $\overline{C}_{week2} = 72.89\% \pm 18.40$, $\overline{C}_{week3} = 78.55\% \pm 15.48$, $\overline{C}_{week4} = 83.68\% \pm 14.23$. Regarding the enjoyment ratings, we found no significant main effects of group ($p = 0.16$), reward ($p = 0.45$) nor interaction effects ($p = 0.25$) (Figure S1, b). Descriptive statistics are as follows: HV: $\overline{E}_{week1} = 56.98\% \pm 18.06$, $\overline{E}_{week2} = 53.15\% \pm 24.72$, $\overline{E}_{week3} = 51.61\% \pm 26.24$, $\overline{E}_{week4} = 53.70\% \pm 30.28$ and OCD: $\overline{E}_{week1} = 59.23\% \pm 15.77$, $\overline{E}_{week2} = 61.19\% \pm 17.78$, $\overline{E}_{week3} = 60.49\% \pm 22.93$, $\overline{E}_{week4} = 64.41\% \pm 26.37$.

### Figure S1

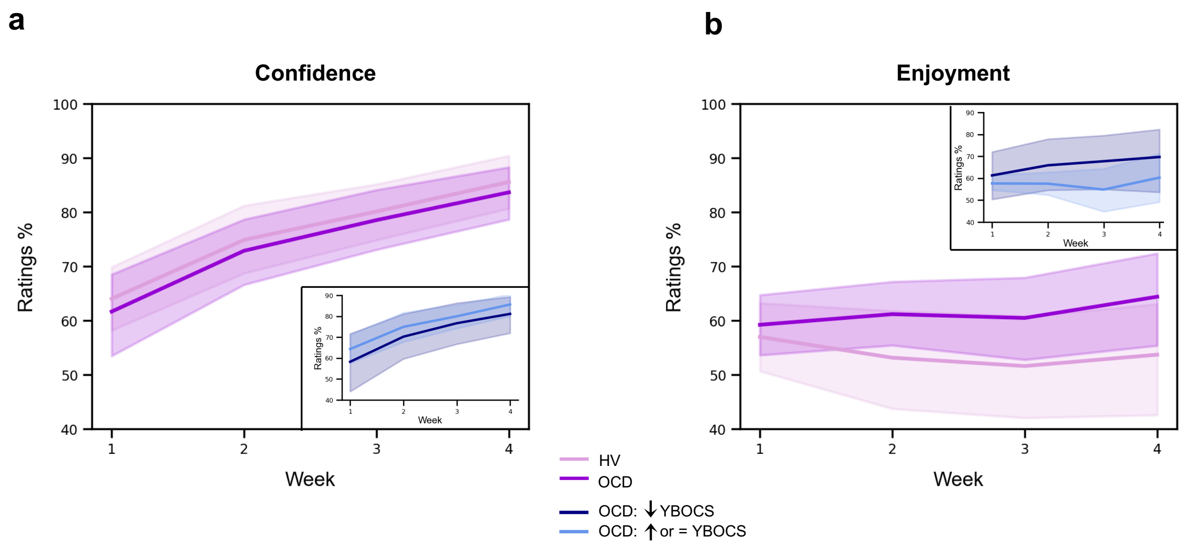


**Figure S1.** Confidence and enjoyment results. The plots depict the average participants' ratings on confidence (a) and enjoyment (b) across the 4 weeks of app training. Solid lines: mean; Transparent regions: confidence interval. Light purple: Healthy Volunteers; Dark purple: patients with OCD. The **insert plots** show the results for the 2 subgroups of the OCD sample, when split based on their YBOCS change after the app training [14 patients with improved symptomatology (reduced YBOCS scores) and 18 patients who remained stable or felt worse (i.e. respectively, unchanged or increased YBOCS scores)].

**Switch session**

Similarly to the learning analysis of the practice sessions, we conducted an individual exponential fitting approach to the switch sessions and assessed between-group differences on performance, both on sequence duration [or movement time (*MT*)] and reaction time (*RT*, i.e. time taken to initiate the sequence, latency). No significant group differences were found in any of the 3 estimated fitting parameters for *MT*: *amount of learning* ($MT_L$): $U = 395$, $p = 0.082$; *learning constant* ($n_r$): $U = 575$, $p = 0.544$ and *asymptote* ($MT_0$): $U = 450$, $p = 0.311$ (Figure S2, a). Descriptive statistics are the following: HV: M $MT_L$ = 1.72 s, $IQR$ = 0.66 s; M $n_r$ = 57, $IQR$ = 50; M $MT_0$ = 1.56 s, $IQR$ = 0.41 s and OCD: M $MT_L$ = 2.16 s, $IQR$ = 1.24 s; M $n_r$ = 49, $IQR$ = 53; M $MT_0$ = 1.67 s, $IQR$ = 0.45 s. Similarly, no significant group differences were found in the 3 estimated fitting parameters for *RT* (Figure S2, b): *reaction time speed-up* achieved over the course of the switch sessions ($R_L$): $U = 470$, $p = 0.45$; *reaction rate* ( $n_R$): $U = 450$, $p = 0.31$ and *reaction time at asymptote* ($R_0$): $U = 552$, $p = 0.75$ (Figure S2, b). Descriptive statistics are the following: HV: M$R_L$ = 1.12 s, $IQR$ = 0.72 s; M $n_R$ = 100, $IQR$ = 168; M $R_0$ = 0.57 s, $IQR$ = 0.50 s and OCD: M$R_L$ = 1.08 s, $IQR$ = 0.70 s; M $n_R$ = 125, $IQR$ = 129; M $R_0$= 0.56 s, $IQR$ = 0.32 s. Since the switching between the two sequences was done in a pseudo-randomised order, we have also assessed *RT* restricting our analysis to the specific trials where the switch occurred (i.e. when sequence 1 was followed by sequence 2 and vice-versa). No group differences in the switching performance were found, meaning that patients with OCD could switch between the two sequences without any difficulties or slowness.

We also investigated potential group differences in accuracy during the switch sessions. After a similar individual exponential fitting approach, statistical analysis of the 3 estimated fitting parameters for Accuracy indicated no group differences (Figure S2, c): *amount of learning* as measured by accuracy achieved over the course of the switch sessions ($Acc_L$): $U = 538$, $p = 0.56$; *learning rate*: $U = 517$, $p = 0.77$ and *asymptote* ($Acc_0$): $U = 419$, $p = 0.29$).

### Figure S2

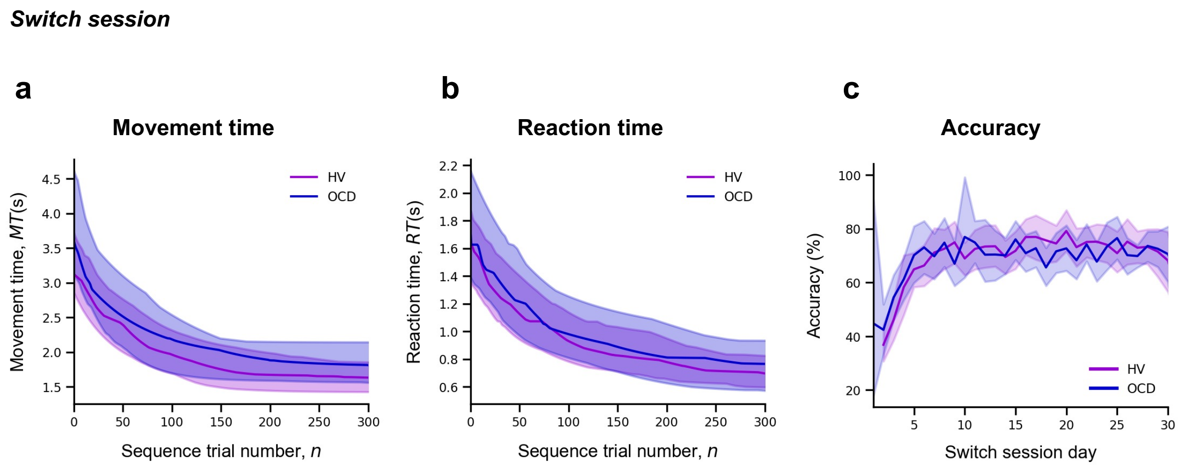


**Figure S2.** Model fitting procedure conducted for the switch sessions. Group comparison resulting from all individual exponential fits modelling the movement time (a), reaction time (b) and accuracy (c) profiles of each participant. No group differences were found. For (a) and (b) plots: solid lines represent median (M) and transparent regions the interquartile range (IQR); For plot (c): solid lines represent the mean and transparent regions for the confidence interval. Purple: healthy volunteers (HV); Blue: patients with OCD.

We did not analyse the short retention speed sessions because no new components were introduced here, as compared to the practice sessions. Therefore, we did not expect any differences between the practice and the speed sessions.

### Extinction

With the goal of promoting habitual actions, the app was designed to remove the explicit reward feedback (points) on the 21st day of practice (extinction procedure). We analyzed the effect of extinction on accuracy, sequence duration (*MT*) and reaction time (*RT*) by comparing the two blocks of practice pre- and post-removal of the rewarding feedback. We analyzed both the practice and switch conditions. While the latter had not previously received reward feedback, it could have potentially been influenced by extinction as a factor. A 2x2 ANOVA with extinction (pre and post extinction) as within-subject factor and group as between-subject factor, indicated no group, extinction or interaction effects neither on accuracy, *RT* or *MT* (Figure S3). Statistic results are as follows: 1) practice sessions; *variable reward*: no effect of group ($p = 0.31$), extinction ($p = 0.28$) or interaction ($p = 0.57$) on accuracy; no effect of group ($p = 0.07$), extinction ($p = 0.99$) or interaction ($p = 0.61$) on *MT*; no effect of group ($p = 0.06$), extinction ($p = 0.53$) or interaction ($p = 0.44$) on reaction time; *continuous reward*: no effect of group ($p = 0.74$), extinction ($p = 0.16$) or interaction ($p = 0.82$) on accuracy; no effect of group ($p = 0.56$), extinction ($p = 0.78$) or interaction ($p = 0.31$) on *MT*; no effect of group ($p = 0.55$), extinction ($p = 0.67$) or interaction ($p = 0.47$) on reaction time; 2) Switch sessions; no effect of group ($p = 0.19$), extinction ($p = 0.17$) or interaction ($p = 0.74$) on accuracy; no effect of group ($p = 0.47$), extinction ($p = 0.89$) or interaction ($p = 0.27$) on *MT*; no effect of group ($p = 0.46$), extinction ($p = 0.78$) or interaction ($p = 0.42$) on reaction time.

### Figure S3

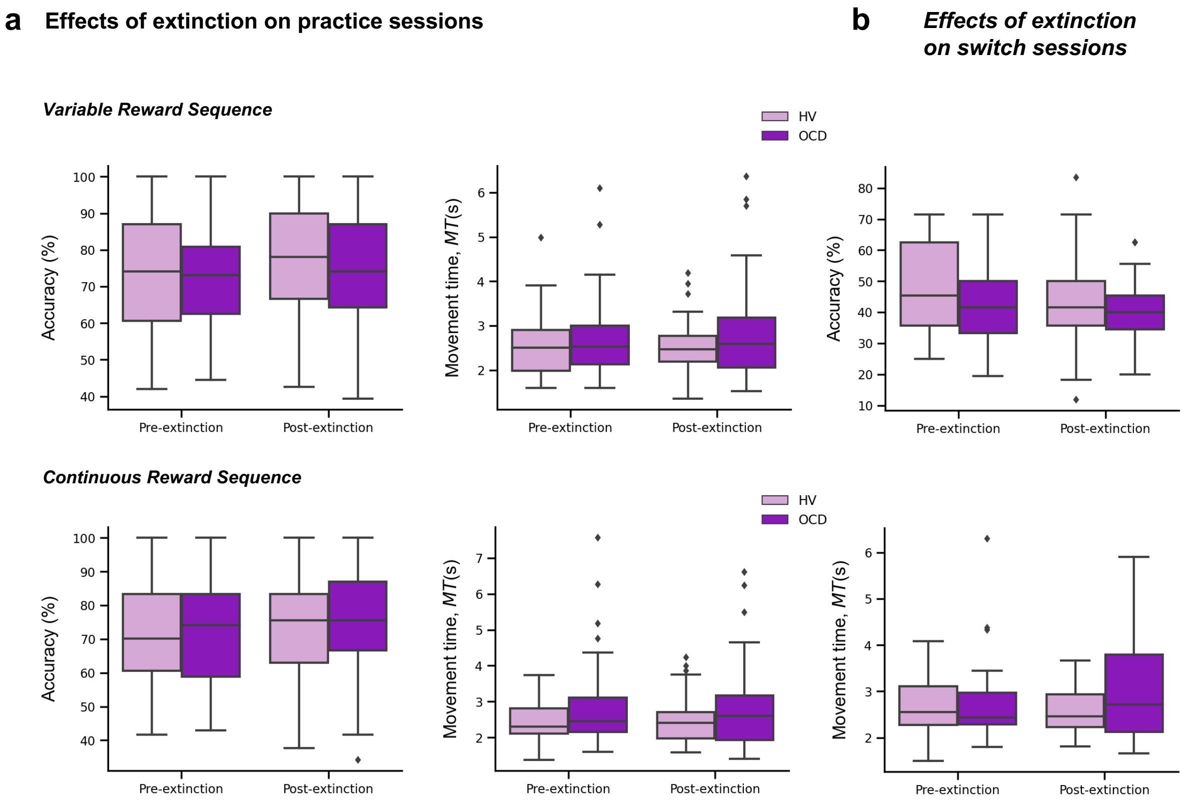


**Figure S3.** Extinction results. a) Effects of extinction on the number of successful trials (accuracy) (*left plots*) and *MT* (*middle plots*) across the practice sessions only, separately for the variable (*upper panel*) and continuous (*lower panel*) reward conditions. b) Effects of extinction on the number of successful trials (accuracy) (*top plot*) and *MT* (*bottom plot*) across the switch sessions. Note that for these analyses we used the two blocks of practice pre- and post-removal of the rewarding feedback (both on the practice and switch conditions).

***Sample size for the reward sensitivity analysis***

The conditional probability distributions $p(\Delta T|\Delta R+)$ and $p(\Delta T\,|\Delta R-)$ were separately fitted to subsamples of the data across continuous reward practices (Figure S1). One practice corresponded to 20 correctly performed sequences. The $p(\Delta T|\Delta R)$ distributions for *MT* *(equation [7])* and the normalized consistency index *normC* were fitted with a non-significantly different number of correct sequences in OCD and in the HV sample (mean 127 [SEM 12] sequences in OCD; 109 [9] sequences in HV; BF < 1/3, moderate evidence against group differences in the number of correct sequences available for this analysis). However, more sequences were available to fit the $p(\Delta T|\Delta R+)$ distribution (mean 107 [standard error of the mean or SEM, 10]) than the $p(\Delta T|\Delta R-)$ distribution (98 [8]). This outcome reflected that participants overall observed more increases than decrements in feedback scores. Importantly, matching both reward and group samples in the number of sequences yielded the same ANOVA results as reported in the main manuscript for the centre and spread of the normalized $\Delta MT$ distribution and *normC* distribution.

Our analysis protocol was designed to ensure that incorrect trials do not contaminate or confound the results. To estimate the trial-to-trial difference in the normalized $\Delta MT$ (or *normC*) and $\Delta R$, we exclusively included pairs of contiguous trials where participants achieved correct performance and received feedback scores for both trials. For example, if a participant made a performance error on trial 23, we did not include $\Delta R$ or $\Delta MT$ estimates for the pairs of trials 23-22 and 24-23. Instead of excluding incorrect trials from our analyses, we retained them in our time series but assigned them a NaN (not a number) value in Matlab®. As a result, $\Delta R$ and $\Delta MT$ was not defined for those two pairs of trials. We followed the same protocol for *normC*. This approach ensured that our analyses are not confounded by incremental or decremental feedback scores between non-contiguous trials. In the past, when assessing the timing of correct actions during skilled sequence performance, we also considered events that were preceded and followed by correct actions. This excluded effects such as post-error slowing from contaminating our results (Bury et al., 2019; Ruiz et al., 2009).

**Figure S4.**

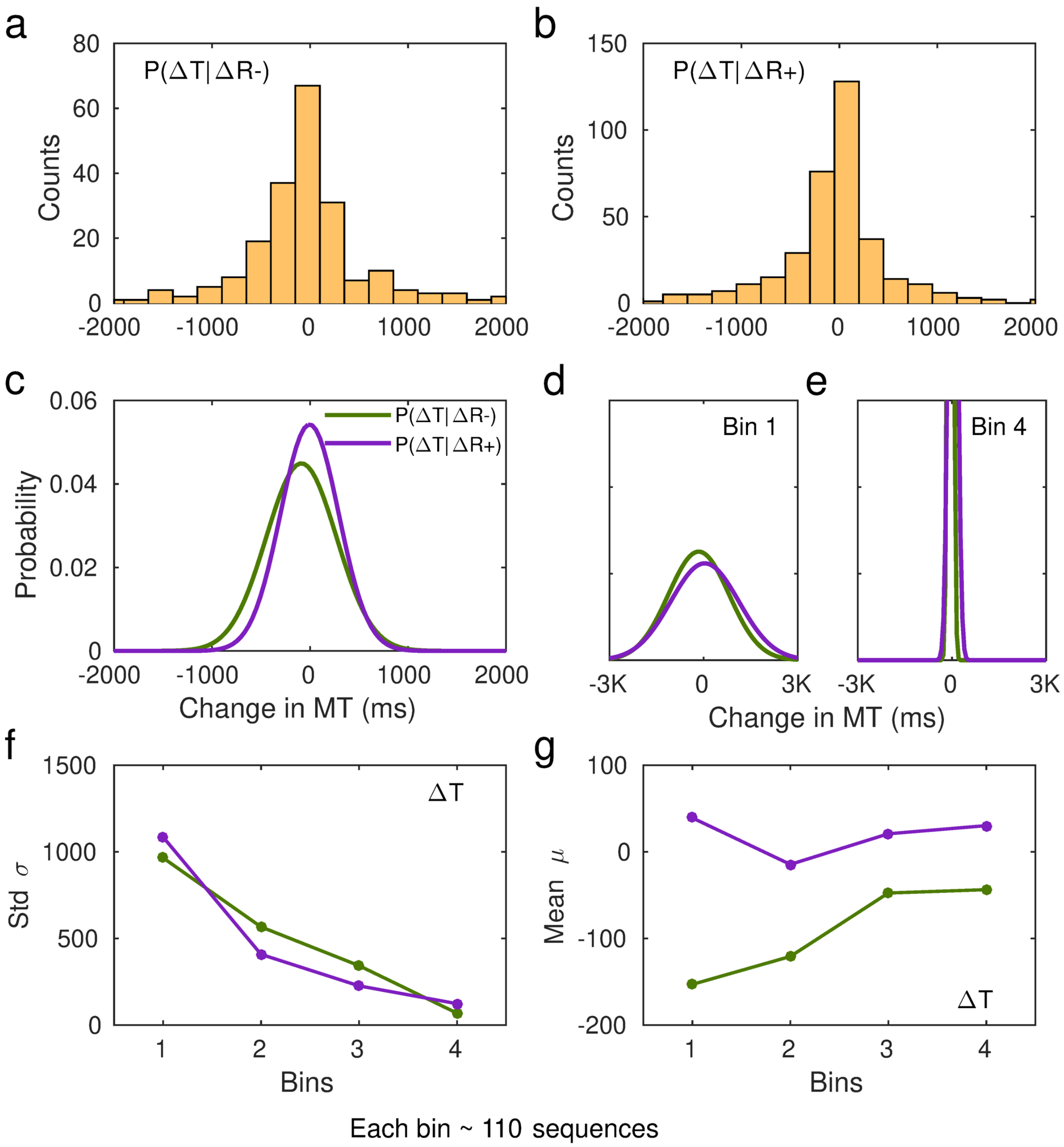


**Figure S4. Fitting of performance data to conditional probability distributions *p(ΔT|ΔR+)* and *p(ΔT |ΔR−)*. a)** Histogram of the changes in non-normalized movement time (*MT*, in ms)

following a decrease in scores relative to the previous trial, $\Delta R-$. Data from one representative subject. **b)** Same as a) but for changes in $MT$ following a reward increment, $\Delta R+$. **c)** Example of fitted Gaussian distributions to the histogram data in a) and b). The Gaussian fit was estimated in MATLAB using the Curve Fitting Toolbox [function *fit*, *fit(x,y,'gauss1')*] on a 20-bin histogram distribution of the data. This example illustrates the Gaussian fits to the total number of sequences and using non-normalized changes in MT (in ms). The full $p(\Delta T|\Delta R+)$ and $p(\Delta T\,|\Delta R-)$ distributions are denoted by the purple and green lines, respectively. **d-e)** In our analyses, we split the total sample of correct sequences that each participant completed over the course of their training into four bins. We then fitted the Gaussian probability distributions to the subsample of sequences in each bin (~110 on average). The first bin is represented in d), while the fourth bin is represented in e). The y-axis limits are identical to those used in panel c). **f-g)** From the fitted Gaussian distributions we obtained the standard deviation, $\sigma$, to assess the spread of the distribution, and the mean, $\mu$. Panel f) displays the std across bins of sequences separately for $p(\Delta T|\Delta R+)$ and $p(\Delta T\,|\Delta R-)$. Panel g) shows the shift in the center of the Gaussian distribution over the course of practiced sequences.

**Figure S5**

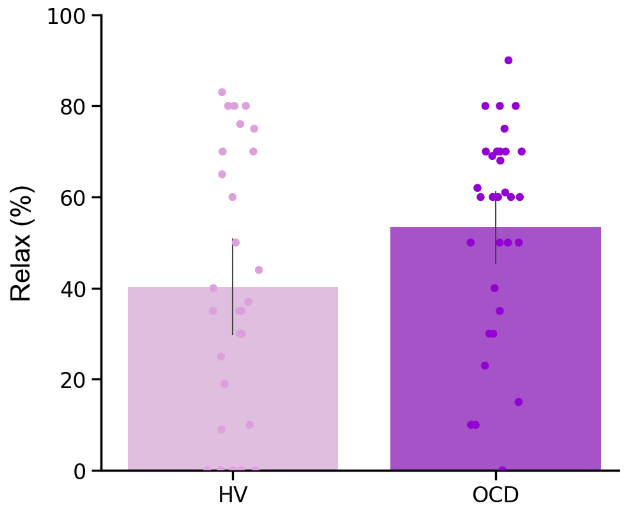

**Figure S5**. **Relaxation ratings**. Participant's ratings on how stressful/relaxing the app training was (rated in a scale from -100% highly stressful to 100% very relaxing. No group differences ($U = 351$, $p = 0.1$).

**Table S4.** Participant's qualitative and quantitative feedback on the impact of the app-training in their life quality

| Population | N | Qualitative feedback | Quantitative feedback (%) |
|---|---|---|---|
| HV | 28 | It was a very neutral game so it did not interfere positively or negatively with my life | 0 |
| | 2 | It did not impact my routines but it was very boring to do this every day | 0 |
| | 1 | I could see the progress and this was fulfilling | 10 |
| | 1 | It kept my brain active in the morning | 15 |
| | 1 | This was a month full of changes (I changed job, moved house, …) so the app was probably the only thing that remain constant throughout the month. I think it probably worked as some kind of mindfulness. | 60 |
| OCD | 16 | It was neutral, it did not interfere with my life in any way | 0 |
| | 4 | The app gave me a goal, some kind of structure to work towards. It was quite relaxing to do it. I enjoyed and repetition was not boring. | 2, 60, 70 and 70 |
| | 1 | It took my mind off the obsessions and rituals. Moreover, playing the app was a challenge to me. Seeing that I was getting better at it gave me a sense of achievement. This increased my confidence, which has spilled over into other areas of my life. | 80 |
| | 1 | I found it quite relaxing and a diversion at times. | 23 |
| | 1 | It was gratifying because I was finally able to complete something | 30 |
| | 1 | It kept me occupied | 10 |
| | 1 | The app made me use my brain and became part of my everyday routine | 60 |
| | 1 | It was definitely a stress relief. I was familiar enough with the app and this was very relaxing. I felt more confident throughout and then this makes me feel really good. I could switch off my obsessions. It was a focus although repetitive. The repetitiveness is relaxing because it is familiar. | 70 |
| | 1 | It took my mind off of other checking rituals for a short time. | 25 |
| | 1 | The app training made me sit and relax and because I had to concentrate I did not worry with other things. | 10 |
| | 1 | It was sometimes a bit of fun while having a difficult day | 30 |
| | 1 | While I was doing I was so focused on doing it that the thoughts were not coming so often. I just don't rate higher because in the last couple of weeks there were some other things in my life that upset me and the old came stronger | 50 |
| | 1 | It gave me sense of achievement | 40 |
| | 1 | Definitely not improved my life. A bit of the opposite as it was an extra thing I needed to do every day | -30 |

Quantitative feedback was given in a scale from -100% to 100%, in which 0 was no change in life quality, -100% was the maximum decrease in life quality and 100% was the maximum increase in life quality
